# Inferring the multiplicity of founder variants initiating HIV-1 infection: a systematic review and individual patient data meta-analysis

**DOI:** 10.1101/2021.07.14.21259809

**Authors:** James Baxter, Sarah Langhorne, Ting Shi, Damien C. Tully, Ch. Julián Villabona-Arenas, Stéphane Hué, Jan Albert, Andrew Leigh Brown, Katherine E. Atkins

**Affiliations:** Usher Institute, The University of Edinburgh, Edinburgh, United Kingdom; Department of Infectious Disease Epidemiology, Faculty of Epidemiology and Population Health, London School of Hygiene and Tropical Medicine, London, United Kingdom; Centre for Mathematical Modelling of Infectious Diseases, London School of Hygiene and Tropical Medicine, London, United Kingdom; Karolinska Institute, Stockholm, Sweden; Institute of Evolutionary Biology, The University of Edinburgh, Edinburgh, United Kingdom

## Abstract

**Background:** HIV-1 infections initiated by multiple founder variants are characterised by a higher viral load and a worse clinical prognosis, yet little is known about the routes of exposure through which transmission of multiple founder variants is most likely.

**Methods:** We conducted a systematic review of studies that estimated founder variant multiplicity in HIV-1 infection, searching MEDLINE, EMBASE and Global Health databases for papers published between 1^st^ January 1990 and 14^th^ September 2020 (PROSPERO study <u>CRD42020202672</u>). Leveraging individual patient estimates from these studies, we performed a logistic meta-regression to estimate the probability that an HIV infection is initiated by multiple founder variants. We calculated a pooled estimate using a random effects model, subsequently stratifying this estimate across nine transmission routes in a univariable analysis. We then extended our model to adjust for different study methods in a multivariable analysis, recalculating estimates across the nine transmission routes.

**Findings:** We included 70 publications in our analysis, comprising 1657 individual patients. Our pooled estimate of the probability that an infection is initiated by multiple founder variants was 0·25 (95% CI: 0·21-0·29), with moderate heterogeneity (*Q* = 132 · 3, *p* < 0 · 001, *I*^2^ = 64 · 2%). Our multivariable analysis uncovered differences in the probability of multiple variant infection by transmission route. Relative to a baseline of male-to-female transmission, the predicted probability for female-to-male multiple variant transmission was significantly lower at 0·13 (95% CI: 0·08-0·20), while the probabilities for people-who-inject-drugs (PWID) and men-who-have-sex-with-men (MSM) transmissions were significantly higher at 0·37 (0·24-0·53) and 0·30 (0·33-0·40), respectively. There was no significant difference in the probability of multiple variant transmission between male-to-female transmission (0·21 (0·14-0·31)), post-partum mother-to-child (0·18 (0·03-0·57)), pre-partum mother-to-child (0·17 (0·08-0·33)), intrapartum mother-to-child (0·27 (0·14-0·40)).

**Interpretation:** We identified PWID and MSM transmissions are significantly more likely to result in an infection initiated by multiple founder variants, whilst female-to-male infections are significantly less likely. Quantifying how the routes of HIV infection impact the transmission of multiple variants allows us to better understand how the evolution and epidemiology of HIV-1 determine clinical outcomes.

**Funding:** This study was supported by the MRC Precision Medicine Doctoral Training Programme (ref: 2259239) and an ERC Starting Grant awarded to KEA (award number 757688). The funding sources played no role in study design, data collection, data analysis, data interpretation, or writing of the report.

**Panel: Research in context:** *Evidence before this study:* Most HIV-1 infections are initiated by a single, genetically homogeneous founder variant. Infections initiated by multiple founders, however, are associated with a significantly faster decline of CD4+ T cells in untreated individuals, ultimately leading to an earlier onset of AIDS. Through our systematic search of MEDLINE, EMBASE and Global Health databases, we identified 82 studies that classify the founder variant multiplicity of early HIV infections. As these studies vary in the methodology used to calculate the number of founder variants, it is difficult to evaluate the multiplicity of founder variants across routes of exposure.

*Added value of this study:* We estimated the probability that an HIV infection is initiated by multiple founder variants across exposure routes, leveraging individual patient data from 70 of the identified studies. Our multivariable meta-regression adjusted for heterogeneity across study methodology and uncovered differences in the probability that an infection is initiated by multiple founder variants by exposure route. While overall, we estimated that 25% of infections are initiated by multiple founder variants, our analysis found that this probability for female-to-male transmission is significantly lower than for male-to-female transmission. By contrast, this probability was significantly higher among people-who-inject-drugs (PWID) and men-who-have-sex-with-men (MSM). There was no difference in the probability of multiple founder variant transmission for mother-to-child transmission when compared with male-to-female sexual transmission.

*Implications of all the available evidence:* Because HIV-1 infections initiated by multiple founders are associated with a poorer prognosis, determining whether the route of exposure affects the probability with which infections are initiated by multiple variants facilitates an improved understanding of how the evolution and epidemiology of HIV-1 determine clinical progression. Our results identify that PWID and MSM transmissions are significantly more likely to result in an infection initiated by multiple founder variants compared to male-to-female. This reiterates the need for focussed public health programmes that reduce the burden of HIV-1 in these risk groups.

## Introduction

Transmission of HIV-1 results in a dramatic reduction in genetic diversity, with a large proportion of infections initiated by a single founder variant.^1,2^ An appreciable minority of infections, however, appear to be the result of multiple founder variants simultaneously initiating infection after a single exposure.^3^ Importantly, these infections caused by multiple founder variants are associated with elevated set point viral load and faster CD4+ T lymphocyte decline.^4–7^

HIV-1 infections initiated via different routes of exposure are subject to different virological, cellular and physiological environments, which likely influence the probability of acquiring infection.^8–10^ For example, the per-act probability of transmission upon exposure is six times and eighteen times higher for transmission between people who inject drugs (PWID) and men who have sex with men (MSM) than for heterosexual transmission.^11^

Despite these differences in the probability of HIV-1 acquisition by route of exposure, there is currently no consensus about whether the route of exposure determines the probability that infection is initiated by multiple founder variants. Differences in selection pressure during transmission have been observed between sexual exposure routes, with less selection occurring during sexual transmission from males to females than vice-versa, and less selection during MSM transmission relative to heterosexual exposure overall.^12,13^ Less selection should lead to more opportunities for infections initiated with more founder variants. Studies quantifying the number of founder variants are, however, inconsistent with these findings, which may be due to differences in methodology and study population.^3,12,14,15^ Moreover, while acquisition risk during sexual transmission is known to be elevated during conditions that increase mucosal inflammation and compromise mucosal integrity, there is no consistent evidence that PWID transmissions, which bypass mucosal barriers altogether, are associated with a higher probability of founder variant initiation.^16,17^ To estimate the role of exposure route on the acquisition of multiple HIV-1 founder variants, we conducted a meta-regression leveraging all available individual patient data, and accounting for heterogeneity across methodology and study population.

## Methods

### Search Strategy and Eligibility Criteria

We searched MEDLINE, EMBASE and Global Health databases for papers published between 1 January 1990 to 14 September 2020 (Appendix S1, ppA2-A6). To be included, studies must have reported original estimates of founder variant multiplicity in people with acute or early HIV-1 infections, be written in English and document ethical approval. Studies were excluded if they did not distinguish between single and multiple founder variants, if they did not detail the methods used, or if the study was conditional on having identified multiple founders. Additionally, studies were excluded if they solely reported data concerning people living with HIV-1 who had known or suspected superinfection, who were documented as having received pre-exposure prophylaxis, or if the transmitting partner was known to be receiving antiretroviral treatment. No restrictions were placed on study design, geographic location, or age of participants. Studies were screened independently by SL and JB. Reviewers were blinded to study authorship during the title and abstract screens, and full text reviews were conducted independently before a consensus was reached; consulting other co-authors when necessary. This review conforms to PRISMA guidelines (Table S2, Appendix ppA7-A10).

### Data Extraction

Individual patient data (IPD) were collated from all studies, with authors contacted if these data were not readily available. Studies were excluded from further analysis if IPD could not be obtained. Only individuals for whom a route of exposure was known were included. Additionally, we removed any entries for individuals with known or suspected superinfection, who were receiving pre-exposure prophylaxis or for whom the transmitting partner was known to be receiving antiretroviral therapy. For the base-case dataset, we recorded whether an infection was initiated by one or multiple variants and eight predetermined covariates to be considered in the multivariable meta-regression:

i. *Route of exposure*. Female-to-male (HSX-FTM), male-to-female (HSX-MTF), men-who-have-sex-with-men (MSM), pre-partum, intrapartum and post-partum mother to child (MTC), or people who inject drugs (PWID).
ii. *Quantification Method.* Methodological groupings were defined by the properties of each approach, resulting in six levels: phylogenetic, haplotype, distance, model, or molecular (Table S1).
iii. *HIV subtype*. Infecting subtypes were classed as either a canonical geographically delimited subtypes (A-D, F-H, J and K), a circulating recombinant form (CRF), or ‘recombinant’ (when a putative recombinant was identified but not designated a CRF).^18,19^
iv. *Delay between infection and sampling*. For sexual or PWID exposures, the delay was classified as either less than or equal to 21 days if the patient was seronegative at time of sampling (Feibig stages I-II) or more than 21 days if the patient was seropositive (Fiebig stages III-VI). For mother-to-child infections, if infection was confirmed at birth, or within 21 days of birth, the delay was classified as either less than or equal to 21 days. A positive mRNA or antibody test reported after this period was classified as a delay of greater than 21 days.
v. *Number of genomes analysed per participant*. For studies that use single genome amplification, this was the number of consensus genomes obtained.
vi. *Genomic region analysed*. The region was classified as envelope (env), pol, gag or near full length genome (NFLG).
vii. *Alignment length analysed*. The length was measured in base pairs, discretised to the nearest 250, 500, 1000, 2000, 4000, 8000, and near full length genome (NFLG) intervals (∼9000).
viii. *Use of single genome amplification (SGA) to generate viral sequences.* A binary classification was used to characterise whether the viral genomic data were generated using SGA. SGA mitigates the risk of Taq-polymerase mediated template switching, nucleotide misincorporation or unequal amplicons resampling encountered in regular bulk or near endpoint polymerase chain reaction (PCR) amplification.^20–22^

If information from any of the covariates iii-viii was missing or could not be inferred from the study, we classified its value as unknown. We excluded covariate levels for which there were fewer than 6 data points. For our main analysis, we removed repeat measurements for the same individual, and used only those from the earliest study or, where the results of different methods were reported by the same study, the conclusive method used for each individual. Further details on covariate selection are in the supplementary methods (Appendix ppA2-A6).

### Statistical Analysis

We calculated a pooled estimate of the probability of multiple founder variant infection using a ‘one-step’ generalised linear mixed model (GLMM); assuming an exact binomial distribution with a normally distributed random effect on the intercept for within-study clustering and fitted by approximate maximum likelihood.^23^ Heterogeneity was measured in terms of *τ*^2^, the between-study variance; I^2^, the percentage of variance attributable to study heterogeneity; and Cochran’s Q, an indicator of larger variation between studies than of subjects within studies.^24^ Publication bias was assessed using funnel plots and Egger’s regression test.^25^ All analyses were conducted in R 4.1.2.^26^

Pooled estimates obtained through a ‘one-step’ approach are usually congruent with the canonical ‘two-step’ meta-analysis model, however discrepancies may arise due to differences in weighting schemes, specification of the intercept or estimation of residual variances.^27^ We compared the results from our ‘one-step’ model with a ‘two-step’ binomial-normal model to confirm our estimates were consistent. We also performed seven sensitivity analyses to test the robustness of our pooled estimate: i) iteratively excluding single studies, ii) excluding studies that contained fewer than ten participants, iii) setting variable thresholds of the number of genomes per patient, iv) excluding studies that consisted solely of single founder infections, v) excluding IPD that did not use single genome amplification, vi) including only those data that matched a ‘gold-standard’ methodology of haplotype-based methods and envelope gene analysis, and vii) an assessment of the effect of vaccine breakthrough, sequencing technologies, and molecular methods. To validate our down-sampling method that used only the most recent study for repeated individual data, we calculated a distribution of pooled estimates by refitting the pooling models to 1000 datasets, each containing one datapoint per individual sampled at random from an individual’s possible measurements.

We extended our ‘one-step’ model by conducting a univariable meta-regression with each covariate contributing a fixed effect and assuming normally distributed random effects of publication. We extended this model to a multivariable analysis. Fixed effects were selected according to a ‘keep it maximal’ principle, in which covariates were only removed to facilitate a non-singular fit and to prevent multicolinearity.^28^ We defined our reference case as heterosexual male-to-female transmission, and evaluated through a gold-standard methodology of haplotype-based methods, analysis of the envelope genomic region and a sampling delay of less than 21 days. We report stratified model estimates of the proportion of infections initiated by multiple founders and bootstrapped 95% confidence intervals across each covariate with all other covariates held at their reference case values. We performed four sensitivity analyses to test the robustness of the selected multivariable meta-regression model: i) iteratively excluding single studies, ii) excluding studies that contained fewer than ten participants, iii) excluding studies that consisted solely of single founder infections, and iv) excluding IPD that did not use single genome amplification. The re-sampling sensitivity analysis was repeated on our selected multivariable model as described above for the univariable model. Further details are in the supplementary methods (Appendix ppA2-A6).

## Results

Our search found 7416 unique papers, of which 7334 were excluded. Of the remaining 380, 207 were excluded after abstract screening, leaving a total of 82 eligible studies for IPD collation (Fig.1).^3,5–7,12,14–17,22,29–102^ We extracted IPD from 80 of these studies, comprising 3251 data points. The 80 selected studies from which IPD were collated, were published between 1992 and 2020. Of the 3251 data points extracted, 1484 were excluded from our base case dataset to avoid repeated measurements; arising either between different studies that analysed the same individuals (resulting in the exclusion of five studies), or from repeat analysis of individuals within the same study. After excluding participants for whom the route of exposure was unknown or for whom one or more of their covariate values pertained to a covariate level that did not meet the minimum number (6) of observations across all participants, the base case dataset for our analysis comprised estimates from 1657 unique patients across 70 studies.

**Figure 1:**
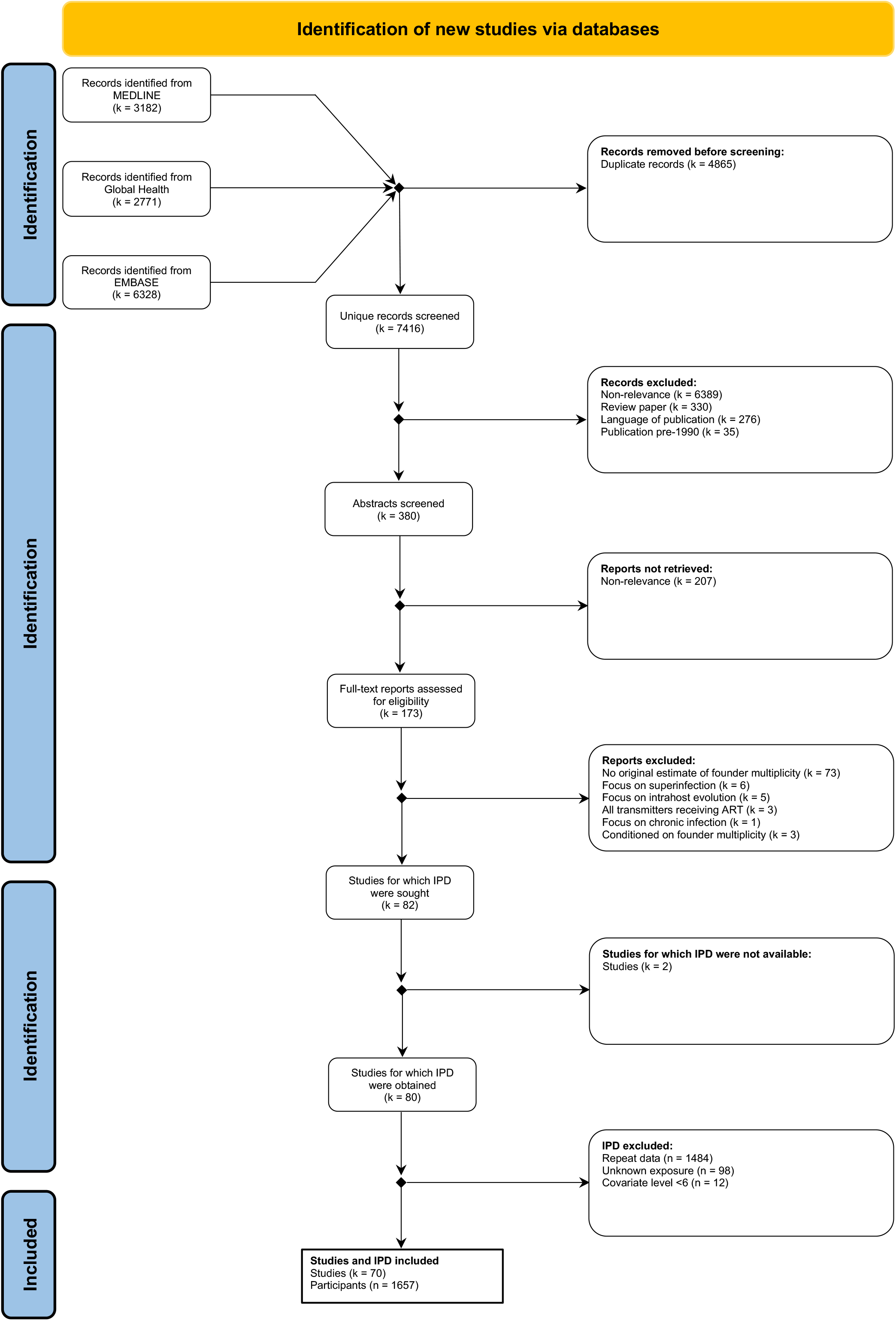
PRISMA flowchart outlining our systematic literature search and the application of exclusion criteria for the individual patient data meta-analysis.

Our base case dataset includes a median of 13 participants per study (range 2-124) and represents infections associated with heterosexual transmission (42·0%, (n=696), MSM transmission (37·4%, n=621), MTC (14·1%, n=234), and PWID transmission (6·4%, n=106) (Fig.2; Table 1; Table S2, Appendix ppA11-17). Among heterosexual transmissions, 67·7% (n=471) were HSX:MTF transmissions, 29·9% (n=208) were HSX:FTM transmissions, with the remainder undisclosed (n=17). Similarly, we subdivided MTC transmission according to the timing of infection with 44·4% (n=104) pre-partum, 24·4% (n=57) intrapartum, 4·7% (n=11) post-partum, with the remainder undisclosed (n=62). Our dataset spanned geographical regions and dominant subtypes, capturing the diversity of the HIV epidemic over time (Fig. S1, Appendix ppA17). Across the base case dataset, 37·1% (n=618) estimates used phylogenetic methods, 26·4% (n=438) used haplotype methods, 20·9% (n=347) used molecular methods, and 13·0% (n=215) and 2·35% (n=39) of estimates were inferred using distance and model-based methods respectively (Fig.2).

**Figure 2:**
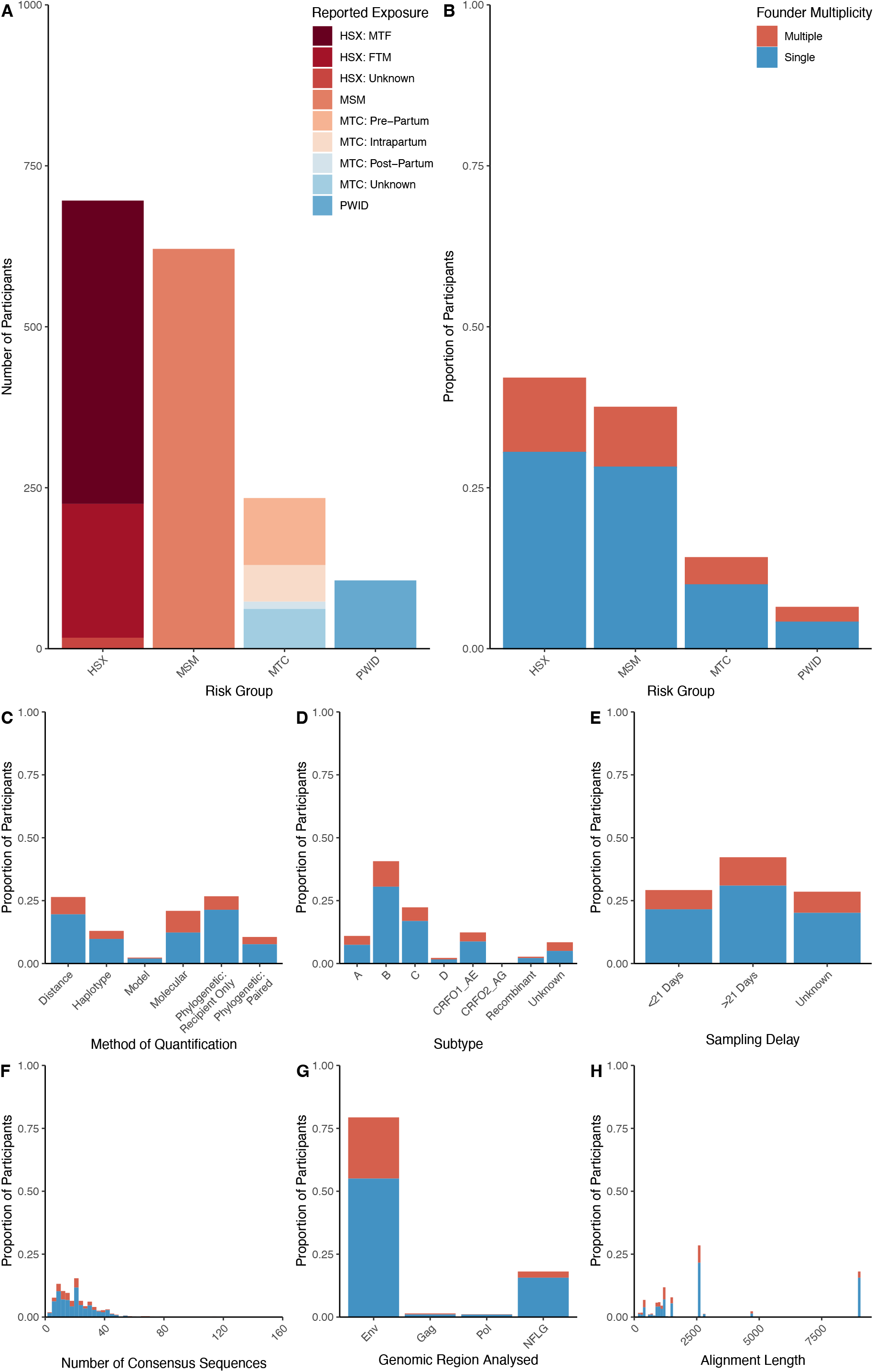
Individual patient data characteristics from the included studies that were tested for inclusion as fixed effects in the multivariable meta-regression model.

**Table 1:** Summary of individual and study characteristics in our base case dataset. Transmission groups recorded as: female-to-male (HSX:FTM), male-to-female (HSX:MTF), men-who-have-sex-with-men (MSM), mother-to-child pre-partum, intrapartum), and post-partum; people who inject drugs (PWID). Numbers within parentheses refer to quantities before removal of repeat participants.

| Transmission Route | Number of studies | Number of participants | Number of participants where multiple founder variant estimated | Quantification methods |  | Genomic regions analysed |  | Number of sequences per participant |
| --- | --- | --- | --- | --- | --- | --- | --- | --- |
| Heterosexual: |  |  |  |  |  |  |  |  |
| MTF | 32 (39) | 471 (601) | 147 (188) | Distance<br>Haplotype<br>Model<br>Molecular<br>Phylogenetic:R<br>Phylogenetic:S&R | 65<br>105<br>9<br>161<br>99<br>32 | Env<br>Gag<br>Pol<br>NFLG | 437<br>14<br>1<br>19 | 21 [2-104] |
| FTM | 25 (30) | 208 (319) | 39 (61) | Distance<br>Haplotype<br>Model<br>Molecular<br>Phylogenetic:R<br>Phylogenetic:S&R | 67<br>73<br>2<br>26<br>25<br>15 | Env<br>Gag<br>Pol<br>NFLG | 179<br>8<br>3<br>18 | 22 [3-80] |
| Unknown | 3 (4) | 17 (22) | 5 (7) | Distance<br>Haplotype<br>Phylogenetic:S&R | 2<br>4<br>11 | Env<br>NFLG | 15<br>2 | 13 [5-27] |
| MSM | 28 (34) | 621 (812) | 154 (205) | Distance<br>Haplotype<br>Model<br>Molecular<br>Phylogenetic:R<br>Phylogenetic:S&R | 80<br>139<br>10<br>27<br>305<br>60 | Env<br>Pol<br>NFLG | 351<br>13<br>257 | 15 [2-149] |
| PWID | 12 (13) | 106 (116) | 38 (45) | Distance<br>Haplotype<br>Model<br>Molecular<br>Phylogenetic:R<br>Phylogenetic:S&R | 1<br>63<br>14<br>9<br>14<br>5 | Env<br>Pol<br>NFLG | 101<br>1<br>4 | 24 [11-163] |
| MTC: |  |  |  |  |  |  |  |  |
| Pre-partum | 7 (7) | 104 (104) | 31 (31) | Model<br>Molecular<br>Phylogenetic:S&R | 2<br>92<br>10 | Env<br>Gag | 103<br>1 | 31 [6-49] |
| Intrapartum | 7 (7) | 57 (57) | 25 (25) | Model<br>Molecular<br>Phylogenetic:S&R | 2<br>32<br>23 | Env | 57 | 17 [6-31] |
| Post-partum | 1 (1) | 11 (11) | 2 (2) | Phylogenetic:S&R | 11 | Env | 11 | .. |
| Unknown | 6 (6) | 62 (62) | 12 (12) | Haplotype<br>Phylogenetic:S&R | 54<br>8 | Env<br>Gag | 61<br>1 | 37 [4-115] |

Our binomial GLMM pooled estimated the probability that an infection is initiated by multiple founder variants as 0·25 (95% CI: 0·21-0·29), identifying moderate heterogeneity (*Q* = 132 · 3, *p* < 0 · 001, *I*^2^ = 64 · 2%). Visual inspection of a funnel plot and a non-significant Egger’s Test (t = - 0·7495, df = 55, p = 0·4568) were consistent with an absence of publication bias (Fig.S9 Appendix ppA26). Sensitivity analyses revealed the pooled estimate was robust to the choice of model, the inclusion of estimates from repeat participants, and to the exclusion of studies that contained fewer than 10 participants (Fig.S2, Appendix ppA19). While restricting the analysis to participants for whom a large (>28) number of sequences were analysed did not change the pooled estimate (0·26 (0·20-0·34)), restricting the analysis to those individuals with fewer than 11 sequences reduced the estimate to 0·21 (0·17-0·25) (Fig.S3 Appendix ppA20). Analysing only data that matched our ‘gold standard’ study methodology slightly increases the pooled estimate (0.28 (95% CI: 0.22-0.35)) (Fig.S3, Appendix ppA20). We did not identify any studies or risk groups that individually influenced the pooled estimate significantly (Fig.S4, Fig.S5 Appendix ppA21-A22). A pooled estimate subgroup analysis of placebo and vaccine participants from studies for which vaccination status was available revealed no discernible influence of trial arm (Fig.S6, Appendix ppA23). Likewise, no discernible difference was identified between sequencing technologies on the pooled estimate (Fig.S7, Appendix ppA24).

We first extended our binomial GLMM with univariable fixed effects. Relative to a reference exposure route of HSX:MTF, we found significantly lower odds of HSX:FTM transmission being initiated by multiple founder variants (Odds Ratio (OR): 0·53 (95% CI 0·33-0·85)), while other exposure routes were not significantly different (Table 2). The univariable analyses also indicated significantly lower odds of identifying multiple founder variants when the near-full-length genome (NFLG) was analysed (OR: 0·38 (95% CI:0·19-0·68)), relative to the envelope genomic region, while molecular methods resulted in significantly greater odds (OR: 1·93 (1·02-3·45)), relative to haplotype methods. NFLG individuals continued to indicate significantly lower odds of identifying multiple founder variants in the absence of individuals analysed using molecular methods (Fig.S8, Appendix ppA25).

**Table 2:** Odds ratios that an HIV-1 infection is initiated by multiple founder variants, inferred from fixed effects coefficients from the univariable and multivariable meta-regression model. Significant effects in bold. MSM - men who have sex with men; PWID - people who inject drugs; NFLG - near full length genome.

|  | Univariable |  | Multivariable |  |
| --- | --- | --- | --- | --- |
|  | Odds Ratio [95% CI] | p-value | Odds Ratio [95% CI] | p-value |
| **Reported Exposure** |  |  |  |  |
| Heterosexual: male-to-female | 1 (reference) | - | 1 (reference) | - |
| Heterosexual: female-to-male | **0.53 [0.33-0.85]** | **0.006** | **0.55 [0.34-0.90]** | **0.011** |
| Heterosexual: undisclosed | 1.81 [0.40-5.91] | 0.340 | 1.72 [0.25-5.24] | 0.364 |
| MSM | 1.33 [0.83-2.03] | 0.238 | **1.61 [1.00-2.34]** | **0.023** |
| Mother-to-child: pre-partum | 1.26 [0.54-2.65] | 0.589 | 0.76 [0.38-1.59] | 0.479 |
| Mother-to-child: intrapartum | 1.87 [0.81-4.02] | 0.148 | 1.34 [0.58-2.86] | 0.461 |
| Mother-to-child: post-partum | 0.73 [0.01-3.56] | 0.772 | 0.79 [0.00-3.58] | 0.794 |
| Mother-to-child: undisclosed | 1.23 [0.45-3.50] | 0.701 | 0.79 [0.29-2.23] | 0.637 |
| PWID | 2.08 [0.91-4.15] | 0.05 | **2.18 [1.17-3.89]** | **0.018** |
| **Quantification Method** |  |  |  |  |
| Haplotype | 1 (reference) | - | 1 (reference) | - |
| Distance | 0.76 [0.35-1.58] | 0.443 | 1.46 [0.76-2.86] | 0.251 |
| Model | 0.53 [0.09-1.39] | 0.265 | **0.36 [0.09-0.87]** | **0.057** |
| Molecular | **1.93 [1.02-3.45]** | **0.026** | **2.05 [1.09-3.53]** | **0.018** |
| Phylogenetic: recipient only | 0.72 [0.42-1.24] | 0.234 | 0.83 [0.48-1.54] | 0.473 |
| Phylogenetic: source & recipient | 0.90 [0.49-1.57] | 0.730 | 0.95 [0.52-1.92] | 0.852 |
| **Genomic Region** |  |  |  |  |
| Envelope | 1 (reference) | - | 1 (reference) | - |
| NFLG | **0.38 [0.19-0.63]** | **0.002** | **0.31 [0.13-0.62]** | **0.000** |
| Gag | 1.13 [0.22-4.51] | 0.857 | 2.14 [0.45-6.80] | 0.220 |
| Pol | 0.31 [0.00-1.36] | 0.171 | 0.31 [0.00-1.13] | 0.155 |
| **Sampling Delay** |  |  |  |  |
| <21 Days | 1 (reference) | - | 1 (reference) | - |
| >21 Days | 1.09 [0.74-1.61] | 0.629 | 1.16 [0.78-1.65] | 0.434 |
| Unknown | 1.42 [0.84-2.72] | 0.201 | 1.39 [0.81-2.43] | 0.220 |

Next, we used a multivariable model to calculate the probability of multiple founder variants across the seven routes of exposure controlling for method, genomic region, and sampling delay (Fig.3, Table 2). A satisfactory fit was confirmed by inspection of binned residuals superimposed over 95% confidence intervals (Fig. S10, Appendix pp A27). Model estimated probabilities were calculated with respect to our ‘gold standard’ methodology. Compared to a HSX:MTF transmission probability of 0·21 (95% CI: 0·14-0·31), we found that HSX:FTM transmissions were less likely to be initiated by multiple founders than male-to-female transmissions, with probability 0·13 (95% CI: 0·08-0·21) (OR: 0·55 (95% CI 0·34-0·88)). Conversely, PWID and MSM transmissions were more likely to be initiated by multiple founders (0·37 (0·24-0·53) and 0·30 (0·22-0·40), respectively), compared to HSX:MTF (OR: 2·18 (1·11-3·89); 1·61 (1·00-2·34)) (Fig. 3A). Stratifying MTC transmissions by the putative timing of infection, we calculated pre-partum exposures were initiated by multiple founders with probability 0·17 (0·08-0·33), post-partum with probability 0·18 (0·03-0·57), and intrapartum transmissions with probability 0·27 (0·14-0·45).

**Figure 3:**
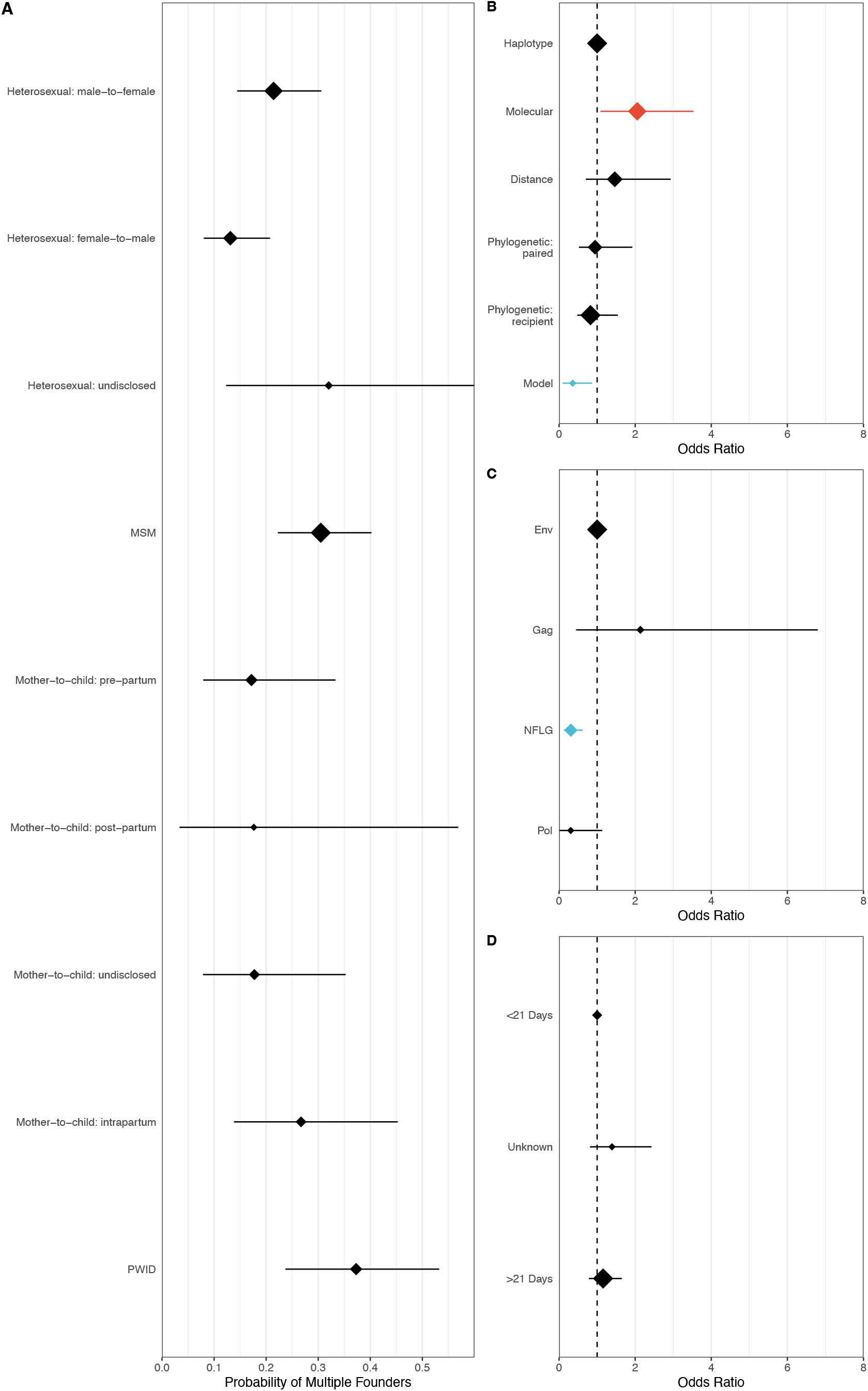
Model estimated probabilities and coefficients obtained from the multivariable model. A) Model estimated probabilities of an infection being initiated by multiple founder variants, stratified by the route of exposure. B-D) Inferred odds ratios of fixed effects variables. Blue denotes that a covariate level significantly decreases the odds of an infection being initiated by multiple founders, whilst red indicates covariate levels for which the odds are significantly greater. For each plot, the reference case is marked at the top of the y axis (dotted line) and the marker size scales with sample size.

We calculated the accuracy of different methods by comparing their estimated probability of multiple founder variants to a gold-standard methodological reference scenario of haplotype-based methods on whole genome sequences with individuals with less than 21 delays between infection and sampling. Our analysis indicates using model-based methods underestimates the chance of multiple founder variants (OR: 0·36 (95% CI: 0·09-0·87)), while using molecular methods results in an overestimation (OR: 2·05 (1·09-3·53)). Compared to the envelope genomic region, analysis of near-full-length genome fragments likely underestimates the proportion of multiple founder infections (OR: 0·31 (95% CI: 0·13-0·62)). Our sensitivity analyses revealed the odds ratios calculated using the uni-and multivariable models are robust to inclusion of data from repeated participants, and to the exclusion of studies that contained fewer than 10 participants, of studies that consisted solely of single founder infections, and of individual data that did not use single genome amplification (Fig.S11, Appendix ppA28).

## Discussion

Using data from 70 published studies, we estimated that a quarter of HIV-1 infections are initiated by multiple founder variants. When controlling for different methodologies across studies, the probability that an infection is initiated by multiple founders decreased from 0·21 (95% CI: 0·14-0·31) for male-to-female infections, to 0·13 (95% CI: 0·08-0·21) for female-to-male infections, but increased for MSM and PWID infections (0·30 (0·22-0·40) and 0·37 (0·24-0·53), respectively). Further, we found that model-based methods, representing a group of approaches that determine founder multiplicity by comparing the observed distribution of diversity with that expected under neutral exponential outgrowth from single variant transmission, were less likely to identify multiple founder infections whereas molecular methods overestimated. Together these results suggest that while the exposure route probably influences the number of founder variants, previous comparison has been difficult due to different study methodologies.

Our pooled estimate is consistent with the seminal study of Keele et al., who found 23·5% (24/102) of their participants had infections initiated by multiple founders.^3^ Our stratified predicted probabilities, however, were marginally higher than those of previous studies. A nine-study meta-analysis of 354 subjects found 0·34 of PWID infections were initiated by multiple founders compared with 0·37 (0·24-0·53) in our study and 0·25 for MSM infections for which we calculated (0·30 (0·22-0·40) ^12^ An earlier meta-analysis of five studies and 235 subjects also found PWID infections were at significantly greater odds than heterosexual infections of being initiated by a single founder, with the frequency of founder variant multiplicity increasing 3-fold, while a smaller, non-significant 1·5-fold increase was observed with respect to MSM transmissions.^16^ In both instances, these studies restricted participants so that the methodology in estimating founder variant multiplicity was consistent. In this study, we were able to leverage individual level data to control for methodological sources of heterogeneity across publications.

Across sexual transmission routes, the probability of multiple founder variants is positively, albeit weakly, associated with an increase in the risk of transmission given exposure. Nonetheless, the probability that infection is initiated by multiple founders remain remarkable consistent. For example, while male-to-male exposures may be up to eighteen times more likely to result in transmission than male-to-female exposures, we calculated a 1·6 fold increase in the risk of multiple founders.^11^ Previously, Thompson et al. reconciled the low probability of acquisition with the relatively high probability of multiple founders by assuming only a fraction of exposures occur in environments conducive for transmission.^103^ In sexual transmission, this could be induced through epithelial damage arising from ulceration or microtrauma; enhancing translocation of viral particles or driving inflammation that propagates recruitment of permissive target cells.^8^ Despite a higher constitutive abundance of permissive cells in the adult human foreskin, the endocervical epithelium and its junction with the ectocervical epithelium are much more susceptible to inflammation and micro-abrasions, reflecting the transmission bias observed in heterosexual transmission.^104,105^

Our analysis has some limitations. First, our classification of founder variant multiplicity is determined by the individual studies, but explicitly defining a founder variant remains challenging. Recent studies have suggested a continuum of genotypic diversity exists, rather than discrete variants, that gives rise to distinct phylogenetic diversification trajectories and may not be reflected by a binary classification.^32,68^ Although a threshold is specified for distance-based methods, this often varies between publications.^106,107^ For example, both Keele et al and Li et al analysed the diversity of the envelope protein, but whilst the former classifies populations with less than 0·47% diversity as homogenous, Li et al included samples up to 0·75%.^3,15^ The distinction between single and multiple founder variants may further be blurred by recombination and hypermutation.^108,109^ Our finding that the analysis of near-full-length genomes were associated with a significant decrease in the odds of multiple founders, suggests earlier studies that rely on smaller, highly variable, fragments of envelop, may have overestimated the frequency of infections initiated by multiple founder variants. Similarly, our sensitivity analyses revealed a subtle correlation between the number of genomes analysed and the probability of observing multiple founder variants, pointing to the possibility that using too few genomes could limit the chance of observing multiple founders.

Second, we acknowledge that some heterogeneity associated with our estimates is encapsulated within the classification of route of exposure. Relying on self-reported route of exposure may bias our results if misclassification occurs systematically across studies. Similarly, insufficient data were available to properly consider risk factors such as genital ulceration, early stage of disease in the transmitter or receptive anal intercourse. These risk factors may confound or mediate any association between the exposure type and the probability of multiple founder variants, potentially hindering a deeper mechanistic understanding as to the risk factors underpinning founder variant multiplicity.^11^ Also, under the hypothesis that the proportion of infections initiated by multiple founders varies by transmission route, our point estimate will be influenced by their relative proportions in our dataset. Globally, it is estimated that 70% of infections are transmitted heterosexually, compared to 42.2% in our dataset.^110^ Our point estimate should be considered a summary of the published data over the course of the HIV-1 epidemic, and not a global estimate at any fixed point in time.

Finally, for several covariates the bootstrapped confidence intervals are wide and may lead to some uncertainty. These are a product of small sample sizes for certain observations, combined with the random effect of publication used in the meta-regression.

This systematic review and meta-analysis demonstrate that infections initiated by multiple founders account for a quarter of HIV-1 infections across major routes of transmission. We find that transmissions involving PWID and MSM are significantly more likely to be initiated by multiple founder variants, whilst HSX:FTM infections are significantly less likely, relative to HSX:MTF infections. Quantifying how the routes of HIV infection impact the transmission of multiple variants allows us to better understand the evolution, epidemiology and clinical picture of HIV transmission.

## Data Availability

Previously published data will be available alongside the code used for this study in a GitHub repository

https://github.com/J-Baxter/foundervariantsHIV_sysreview

## Contributors

KEA conceived the study. JB, SL, DT, KEA designed the study. JB and SL extracted the data. JB performed the experiments and analysed the data. JB and KEA verified the data. All authors interpreted the data. JB and KEA drafted the manuscript, with critical revisions from all authors. All authors had full access to all the data in the study and had final responsibility for the decision to submit for publication.

## Data Sharing

Code and individual patient data used in this study is publicly available at https://github.com/J-Baxter/foundervariantsHIV_sysreview.

## Acknowledgements

JB was supported by the MRC Precision Medicine Doctoral Training Programme (ref: 2259239); CJV-A and KEA were funded by an ERC Starting Grant (award number 757688) awarded to KEA. We are grateful to Morgane Rolland for agreeing to share additional individual patient data with the authors to complete this study. We thank the four anonymous reviewers for their helpful feedback.

## Appendices

### Supplementary Methods

#### Protocol Registration

This systematic review and meta-analysis was registered with PROSPERO following the initial literature search (PROSPERO study <u>CRD42020202672</u>).

#### Full search query submitted to MEDLINE, EMBASE and Global Health databases

(((((transmi*.af. or found*.af. or bottleneck.af. or single.af. or multiple.af. or multiplicity.af. or breakthrough.ti. or TF.af.) and (virus*.af. or variant*.af. or strain.af. or lineage.af. or phenotyp*.af.)) and (HIV.ti. or HIV-1.ti. or human immunodeficiency virus.ti. or env.ti. or envelope.ti or gag.ti. or pol.ti.)) and ((single genome amplification.af. or sga.af. or sgs.af. or ((sequencing.af. or characterized.af.) and (single genome.af. or deep.af. or whole genome.af. or full length.af. or full-length.af.))) or divers*.af. or distance.af. or poisson-fitter.af. or fitness.af. or (monophyletic.af. or paraphyletic.af. or polyphyletic.af.) or (phylogenetic*.af. and (clade.af. or topology.af. or tree.af. or linked.af. or diver*.af. or distance.af. or sieve.af. or molecular dating.af.)))) not ((SIV.ti,ab. or simian immunodeficiency.ti,ab. or fiv.ti,ab. or feline immunodeficiency virus.ti,ab. or exp Hepacivirus/ or Hepatitis.ti,ab. or exp Flaviviridae/ or Tuberculosis.ti,ab. or Enterovirus.ti,ab. or exp Spumavirus/ or diarrhoea.ti,ab. or diarrhea.ti,ab. or superinfection.ti. or exp Malaria/ or CMV.ti,ab. or HPV.ti,ab. or SHIV.ti,ab. OR exp HIV-2/ or phylogeo*.af. or network.ti. or exp HIV Protease Inhibitors/ or exp HIV Integrase Inhibitors/)))

Databases Queried:

- Ovid MEDLINE(R) and Epub Ahead of Print, In-Process & Other Non-Indexed Citations, Daily and Versions(R)
- Global Health 1910 to 2020 Week 36
- EMBASE & EMBASE Classic 1947 – Sep 11

#### Data Extraction

- Route of exposure

We used the route of exposure of horizontally transmitted infections as reported by the original studies. These data are typically ascertained from risk behaviour questionnaires or enrolment criteria for a study cohort. In the majority of cases, a single route of exposure was reported. We stratified the route of exposure as much as possible, given the data available. This resulted in the following levels being included in the models: Female-to-male (HSX-FTM), male-to-female (HSX-MTF), men-who-have-sex-with-men (MSM), pre-partum, intrapartum and post-partum mother to child (MTC), and people who inject drugs (PWID)).

We refer to these as routes of exposure, rather than transmission route as an element of uncertainty is always present; both because there can be multiple concurrent routes of exposure and more generally because self-reported exposure does not necessarily match with transmission. For the same reason we chose not to stratify sexual exposure route into receptive vs. insertive anal sex for male-to-male and vaginal vs. anal sex for male-to-female.

We categorised mother-to-child (MTC) infections into pre-partum, intrapartum and post-partum was using the criteria below. Where these data were nor reported or the results ambiguous, we used the category ‘unknown timing’ (Bertolli et al., 1996):

| Timing | HIV RNA+/PCR+ |
| --- | --- |
| Pre-partum | Infant tests positive at birth |
| Intrapartum | Infant tests negative at birth, but later tests positive after no more than 3 months |
| Post-partum | Infant tests negative at birth, but later tests positive after more than 3 months |

- Sampling Delay

For horizontally transmitted infections, the delay between infection and sampling (not diagnosis) was determined according to seropositivity. A delay of less than or equal to 21 days was recorded if the patient was seronegative at time of sampling (Feibig stages I-II) or more than 21 days if the patient was seropositive (Fiebig stages III-VI). For vertical transmissions, if infection was confirmed at birth, or within 21 days of birth, the delay was classified as either less than or equal to 21 days. A positive mRNA or antibody test definitively reported after this period was classified as a delay of greater than 21 days.

- Methodologies

A binary classification (yes or no) was inferred as to whether the viral genomic data were generated using SGA. Regular bulk or near endpoint polymerase chain reaction (PCR) amplification can generate significant errors such as Taq-polymerase mediated template switching, nucleotide misincorporation or unequal amplicons resampling. (Meyerhans et al. 1990, Simmonds et al. 1990). In SGA, serial dilutions of viral nucleic acids are made, which, assuming the proportion of positive PCR reaction at each dilution follows a null Poisson distribution, reduces the final reactions to contain a single variant that can be cloned, sequenced, and then analysed. (Simmonds et al. 1990, Salazar-Gonzalez et al. 2008).

Quantification method groupings were defined by the properties of each approach, resulting in six levels: phylogenetic, haplotype, distance, model, or molecular (Table S1). Prior the widespread application of sequencing, molecular methods such as heteroduplex mobility assays could provide a qualitative measure of diversity (Novitsky et al. 1996). Heterogeneous genomic segments would form heteroduplexes during gel electrophoresis of viral RNA, allowing one to distinguish genetically similar and dissimilar segments. Although estimates derived from these assays were regarded as close approximations of viral diversity, they only consider a tiny fraction of the whole genome and cannot provide further information regarding phylogenetics, or functional attributes of any substitutions. As a result they may lead to overestimation of the number of founder variants initiating infection.

Distance and model-based methods assume a threshold or distribution of diversity that is reasonably expected to occur under a hypothesis of neutral exponential growth from a single founder and determine whether the observed diversity is consistent with the modelled values (Slatkin and Hudson, 1991; Lee et al., 2009). Within the model category, we include any mathematical or statistical model which tests whether the observed patterns of diversity can be explained by the transmission of a single variant. For example, this includes Poissonfitter, where frequency distributions of Hamming Distances that significantly diverge from the expected Poisson distribution, after controlling for APOBEC mediated hypermutation, represent an over-dispersed population (Giorgi et al., 2010); simple probabilistic models expressing the expected number of substitutions, and estimates of the time to most recent common ancestor that do not involve the reconstruction of genealogies.

**Table S1:** Methods of quantification. Groupings of methods used to infer the founder variant multiplicity of HIV-1 infections. Model and phylogenetic methods may present as similar metrics such as the most recent common ancestor (tMRCA) and topology, but model-based approaches, unlike phylogenetic methods, do not use genealogical information in their calculation and instead are statistical models applied directly to the genomic data.

| Molecular | Haplotype | Distance | Model | Phylogenetic |  |
| --- | --- | --- | --- | --- | --- |
|  |  |  |  | Recipient Only | Source & Recipient |
| Heteroduplex Assay | Highlighter plot<br>Haplotype Frequency | Pairwise distance<br>Diversity | Poissonfitter<br>• Goodness of fit<br>• Starlike topology<br>• tMRCA | Starlike topology<br>tMRCA (genealogy)<br>Diversification | Paired topologies<br>tMRCA (genealogy) |

Haplotype methods identify linkage patterns of individual polymorphisms across samples from a patient. In the study of HIV founder infection multiplicity, this category mostly concerns the use of highlighter plots, that visually map nucleotide mismatches along an aligned gene segment (Keele et al. 2008). Inspection of these graphs facilitates an approximate enumeration of the number of variants initiating an infection and allow for inference of putative recombinants and APOBEC mediated hypermutation, which would erroneously inflate diversity measures. Haplotype methods may also refer to modelling the distribution of haplotypes obtained through longitudinal deep-sequence samples.

Phylogenetic methods are here defined as approaches that explicitly reconstruct ancestral genealogical relationships directly from sequence data. These either use recipient sequences only, in which case a star-like topology is expected to be observed for single founder infections or use source and recipient sequences from known transmission pairs, such that the number of distinct clades of recipient sequences nested within the source sequences corresponds to the number of founder variants.

#### Statistical Models

- Pooled estimates models

We assumed a binary outcome *y_ik_* of whether the infection of individual *k* of study *i* was initiated by multiple founder variants (1) or not (0) with probability (*y_ik_* For the two-step model, we first fit a logit model to these binary outcome data for each study, *i*, where *θ_i_* is the effect size of study *i*, *x_ik_* is whether the infection of individual *k* of study *i* was initiated by multiple founder variants (1) or not (0) and *a_i_* is the intercept :

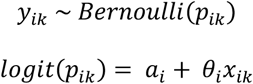

We then accounted for between study variation in the effect sizes by assuming a random effects model such that each of the estimated study effect sizes, 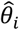, is a sample from a different normal distribution with a mean equal to an underlying study-specific effect size. This study-specific effect size is itself drawn from a normal distribution with constant mean and variance, *τ*^2^ (the between-study variance) :

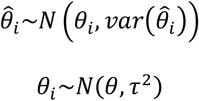

For the one-step model, individual-level and study-level variation are considered simultaneously:

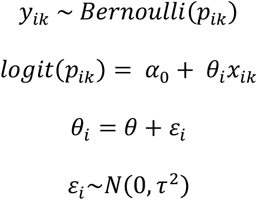

Here, *α*_0_ represents a fixed intercept and *θ_i_* is the random effect of study acting on observation *x_ik_*. *θ_i_* is the sum of *θ*, the mean study effect size, and *ε_i_*, the study-specific effect drawn from a normal distribution with variance, *τ*^2^ (the between-study variance). We compared the results from our one-step model with a two-step model to confirm our estimates were consistent.

- Univariable and multivariable models

We extended our one-step model by conducting a univariable meta-regression with each covariate contributing a fixed effect and assuming normally distributed random effects of publication. We report results for univariable models that analysed the role of route of exposure, quantification method, genome region analysed and sampling delay as fixed effects (*β_ik_*).

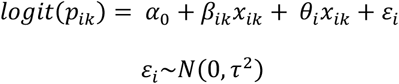

A multivariable model was built from the fixed effects used in the univariable analysis. The fixed effects (*β_nik_*, *n* ∈ [1, *m*]) were selected according to a ‘keep it maximal’ principle, in which covariates were only removed to facilitate a non-singular fit. The selected model is outlined here:

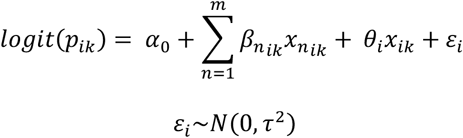

The selected model was assessed for convergence, singularity, multicollinearity using the R package ggeffects. We calculated the proportion of binned residuals within 95% confidence limits. Model estimated probabilities per transmission route were calculated controlling baseline covariates as our ‘gold standard’ methodology (envelope genomic region, a short delay, haplotype analysis).

#### Software and Computational Methods

- All code associated with this study is available under GNU General Public License v3.0 at the following GitHub repository: <u>foundervariantsHIV_sysreview</u>. Further details on how to run the analysis are included in the README.md.

The analyses were conducted in R 4.1.2, principally using the following packages:

lme4, 1.1-27.1, (Bates et al. 2007)
metafor, 3.0-2, (Viechtbauer 2010)
tidyverse, 1.3.1, (including ggplot2 3.3.5, stringr 1.4.0, forcats 0.5.1 & dplyr 1.0.7) (Wickham et al., 2019)
reshape2 1.4.4 (Wickham, 2012)
ggeffects 1.1.1 (Lüdecke, 2018)
mltools 0.5.2
parallel 3.6.2

### Prisma Checklist

**Table S2:** PRISMA checklist referencing the necessary steps taken to pages in this manuscript.

| PRISMA-IPD<br>Section/topic | Item<br>No | Checklist item | Reported<br>on page |
| --- | --- | --- | --- |
| Title |  |  |  |
| Title | 1 | Identify the report as a systematic review and meta-analysis of individual participant data. | 1 |
| Abstract |  |  |  |
| Structured<br>summary | 2 | Provide a structured summary including as applicable: | 2 |
|  |  | **Background:** state research question and main objectives, with information on participants, interventions, comparators and outcomes. |  |
|  |  | **Methods:** report eligibility criteria; data sources including dates of last bibliographic search or elicitation, noting that IPD were sought; methods of assessing risk of bias. |  |
|  |  | **Results:** provide number and type of studies and participants identified and number (%) obtained; summary effect estimates for main outcomes (benefits and harms) with confidence intervals and measures of statistical heterogeneity. Describe the direction and size of summary effects in terms meaningful to those who would put findings into practice. |  |
|  |  | **Discussion:** state main strengths and limitations of the evidence, general interpretation of the results and any important implications. |  |
|  |  | **Other:** report primary funding source, registration number and registry name for the systematic review and IPD meta-analysis. |  |
| Introduction |  |  |  |
| Rationale | 3 | Describe the rationale for the review in the context of what is already known. | 4 |
| Objectives | 4 | Provide an explicit statement of the questions being addressed with reference, as applicable, to participants, interventions, comparisons, outcomes and study design (PICOS). Include any hypotheses that relate to particular types of participant-level subgroups. | 4 |
| Methods |  |  |  |
| Protocol and<br>registration | 5 | Indicate if a protocol exists and where it can be accessed. If available, provide registration information including registration number and registry name. Provide publication details, if applicable. | 2, A2 |
| Eligibility criteria | 6 | Specify inclusion and exclusion criteria including those relating to participants, interventions, comparisons, outcomes, study design and characteristics (e.g. years when conducted, required minimum follow-up). Note whether these were applied at the study or individual level i.e. whether eligible participants were included (and ineligible participants excluded) from a study that included a wider population than specified by the review inclusion criteria. The rationale for criteria should be stated. | 4 |
| Identifying studies - information sources | 7 | Describe all methods of identifying published and unpublished studies including, as applicable: which bibliographic databases were searched with dates of coverage; details of any hand searching including of conference proceedings; use of study registers and agency or company databases; contact with the original research team and experts in the field; open adverts and surveys. Give the date of last search or elicitation. | 4 |
| Identifying studies - search | 8 | Present the full electronic search strategy for at least one database, including any limits used, such that it could be repeated. | A2 |
| Study selection processes | 9 | State the process for determining which studies were eligible for inclusion. | 4 |
| Data collection processes | 10 | Describe how IPD were requested, collected and managed, including any processes for querying and confirming data with investigators. If IPD were not sought from any eligible study, the reason for this should be stated (for each such study). | 4 |
|  |  | If applicable, describe how any studies for which IPD were not available were dealt with. This should include whether, how and what aggregate data were sought or extracted from study reports and publications (such as extracting data independently in duplicate) and any processes for obtaining and confirming these data with investigators. |  |
| Data items | 11 | Describe how the information and variables to be collected were chosen. List and define all study level and participant level data that were sought, including baseline and follow-up information. If applicable, describe methods of standardising or translating variables within the IPD datasets to ensure common scales or measurements across studies. | 5, A2-A4 |
| IPD integrity | A1 | Describe what aspects of IPD were subject to data checking (such as sequence generation, data consistency and completeness, baseline imbalance) and how this was done. | 5 |
| Risk of bias assessment in individual studies. | 12 | Describe methods used to assess risk of bias in the individual studies and whether this was applied separately for each outcome. If applicable, describe how findings of IPD checking were used to inform the assessment. Report if and how risk of bias assessment was used in any data synthesis. | 5 |
| Specification of outcomes and effect measures | 13 | State all treatment comparisons of interests. State all outcomes addressed and define them in detail. State whether they were pre-specified for the review and, if applicable, whether they were primary/main or secondary/additional outcomes. Give the principal measures of effect (such as risk ratio, hazard ratio, difference in means) used for each outcome. | 5 |
| Synthesis methods | 14 | Describe the meta-analysis methods used to synthesise IPD. Specify any statistical methods and models used. Issues should include (but are not restricted to):<br>· Use of a one-stage or two-stage approach.<br>· How effect estimates were generated separately within each study and combined across studies (where applicable).<br>· Specification of one-stage models (where applicable) including how clustering of patients within studies was accounted for.<br>· Use of fixed or random effects models and any other model assumptions, such as proportional hazards.<br>· How (summary) survival curves were generated (where applicable).<br>· Methods for quantifying statistical heterogeneity (such as $I^2$ and $t^2$).<br>· How studies providing IPD and not providing IPD were analysed together (where applicable).<br>· How missing data within the IPD were dealt with (where applicable). | 6, A4-A6 |
| Exploration of variation in effects | A2 | If applicable, describe any methods used to explore variation in effects by study or participant level characteristics (such as estimation of interactions between effect and covariates). State all participant-level characteristics that were analysed as potential effect modifiers, and whether these were pre-specified. | 6 |
| Risk of bias across studies | 15 | Specify any assessment of risk of bias relating to the accumulated body of evidence, including any pertaining to not obtaining IPD for particular studies, outcomes or other variables. | NA |
| Additional analyses | 16 | Describe methods of any additional analyses, including sensitivity analyses. State which of these were pre-specified. | 6 |
| **Results** |  |  |  |
| Study selection and IPD obtained | 17 | Give numbers of studies screened, assessed for eligibility, and included in the systematic review with reasons for exclusions at each stage. Indicate the number of studies and participants for which IPD were sought and for which IPD were obtained. For those studies where IPD were not available, give the numbers of studies and participants for which aggregate data were available. Report reasons for non-availability of IPD. Include a flow diagram. | 9 |
| Study characteristics | 18 | For each study, present information on key study and participant characteristics (such as description of interventions, numbers of participants, demographic data, unavailability of outcomes, funding source, and if applicable duration of follow-up). Provide (main) citations for each study. Where applicable, also report similar study characteristics for any studies not providing IPD. | 9-11 |
| IPD integrity | A3 | Report any important issues identified in checking IPD or state that there were none. | NA |
| Risk of bias within studies | 19 | Present data on risk of bias assessments. If applicable, describe whether data checking led to the up-weighting or down-weighting of these assessments. Consider how any potential bias impacts on the robustness of meta-analysis conclusions. | A18-A27 |
| Results of individual studies | 20 | For each comparison and for each main outcome (benefit or harm), for each individual study report the number of eligible participants for which data were obtained and show simple summary data for each intervention group (including, where applicable, the number of events), effect estimates and confidence intervals. These may be tabulated or included on a forest plot. | 10, A11-A17 |
| Results of syntheses | 21 | Present summary effects for each meta-analysis undertaken, including confidence intervals and measures of statistical heterogeneity. State whether the analysis was pre-specified, and report the numbers of studies and participants and, where applicable, the number of events on which it is based. | 12-14 |
|  |  | When exploring variation in effects due to patient or study characteristics, present summary interaction estimates for each characteristic examined, including confidence intervals and measures of statistical heterogeneity. State whether the analysis was pre-specified. State whether any interaction is consistent across trials. |  |
|  |  | Provide a description of the direction and size of effect in terms meaningful to those who would put findings into practice. |  |
| Risk of bias across studies | 22 | Present results of any assessment of risk of bias relating to the accumulated body of evidence, including any pertaining to the availability and representativeness of available studies, outcomes or other variables. | NA |
| Additional analyses | 23 | Give results of any additional analyses (e.g. sensitivity analyses). If applicable, this should also include any analyses that incorporate aggregate data for studies that do not have IPD. If applicable, summarise the main meta-analysis results following the inclusion or exclusion of studies for which IPD were not available. | A18-A27 |
| **Discussion** |  |  |  |
| Summary of evidence | 24 | Summarise the main findings, including the strength of evidence for each main outcome. | 15 |
| Strengths and limitations | 25 | Discuss any important strengths and limitations of the evidence including the benefits of access to IPD and any limitations arising from IPD that were not available. | 16 |
| Conclusions | 26 | Provide a general interpretation of the findings in the context of other evidence. | 15-16 |
| Implications | A4 | Consider relevance to key groups (such as policy makers, service providers and service users). Consider implications for future research. | 16 |
| **Funding** |  |  |  |
| Funding | 27 | Describe sources of funding and other support (such as supply of IPD), and the role in the systematic review of those providing such support. | 2, 7 |

### Table of Selected Studies

**Table S3:**
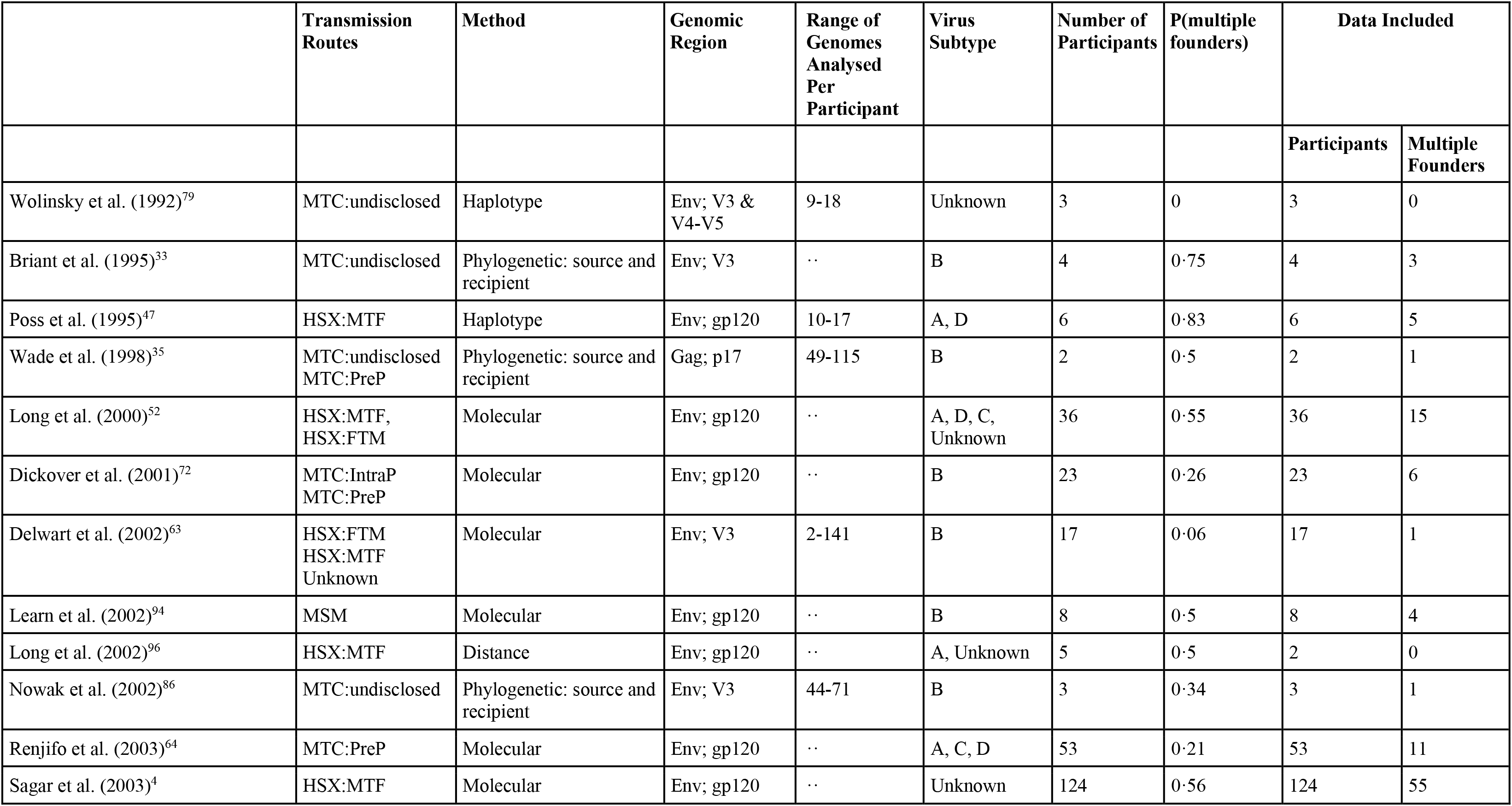
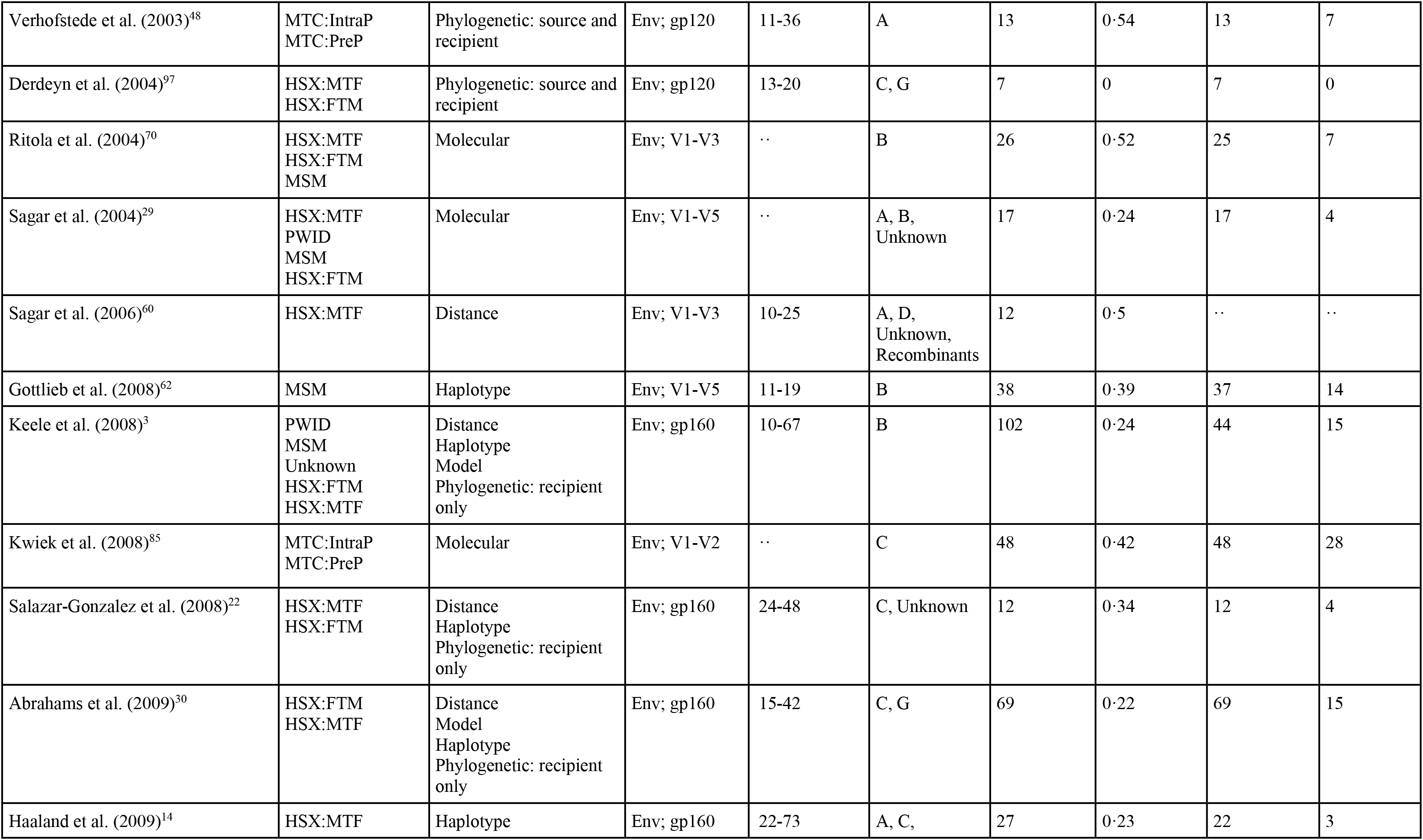
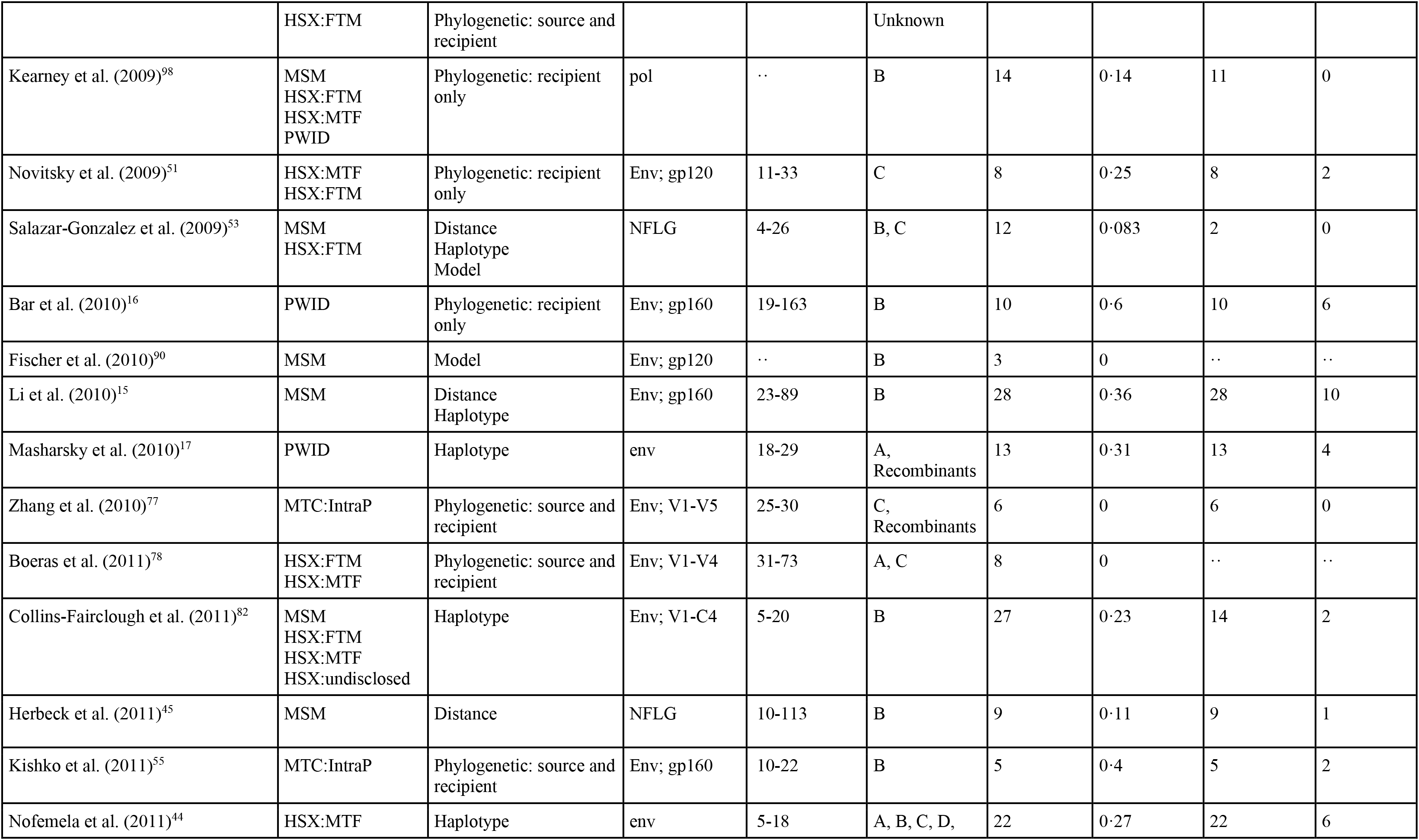
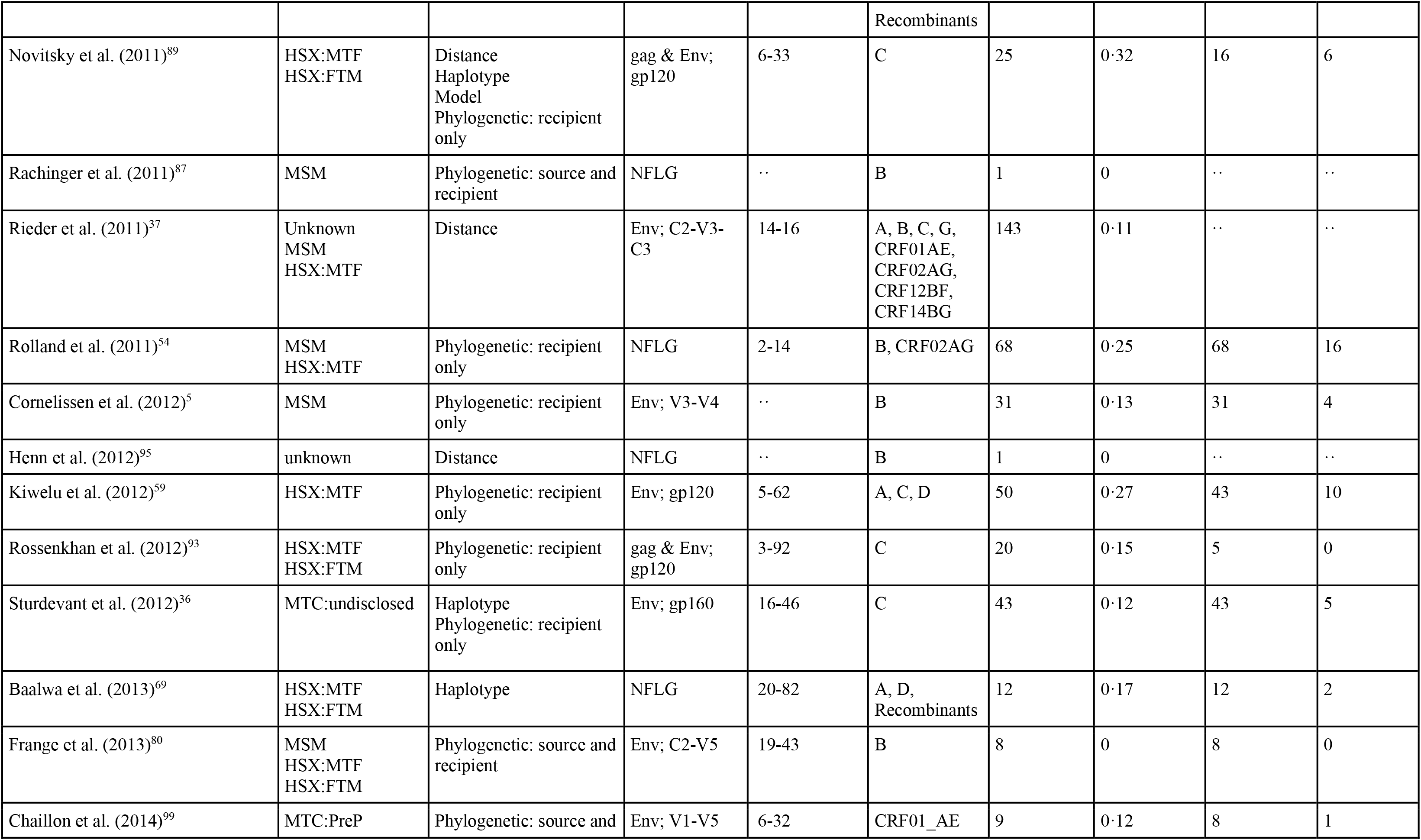
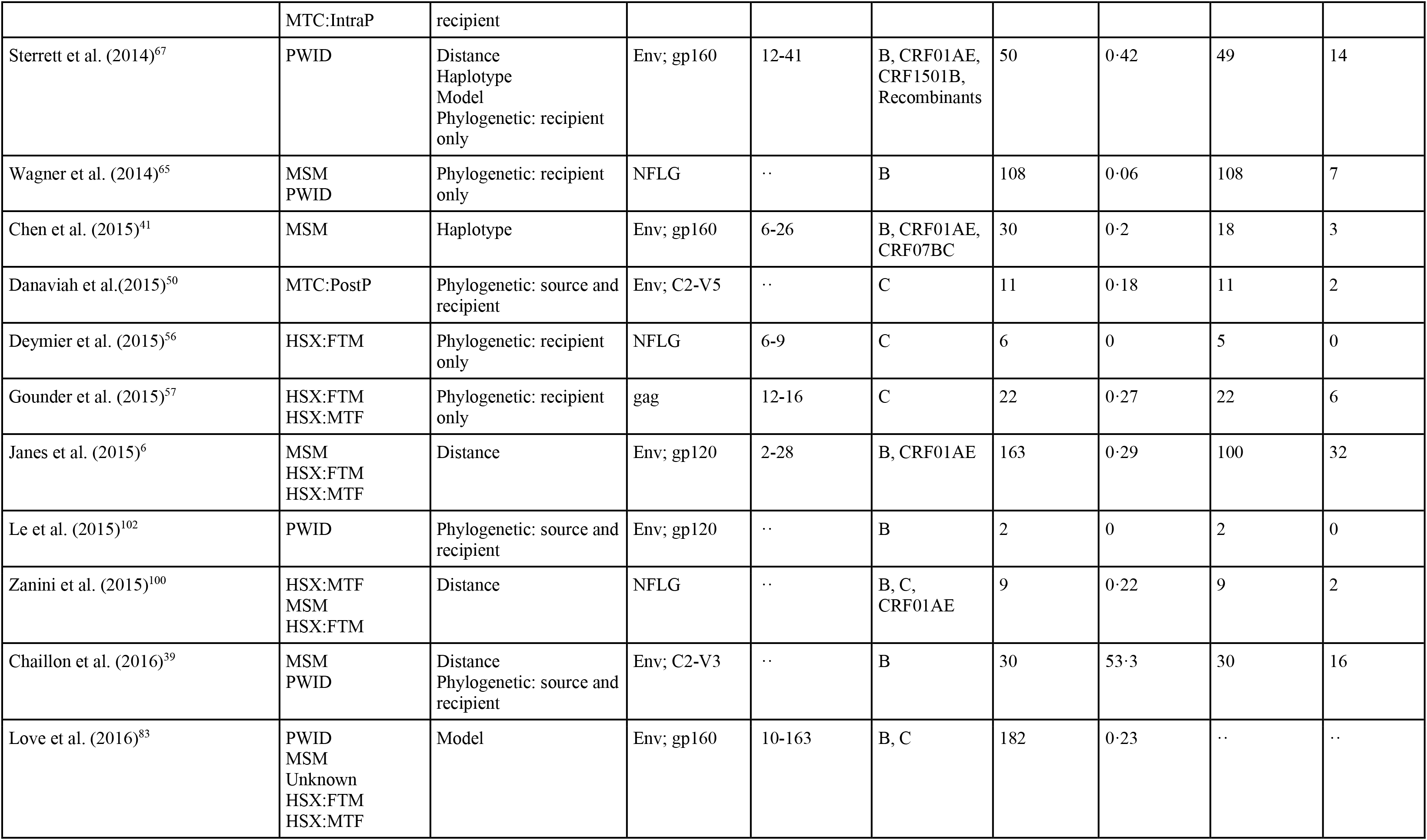
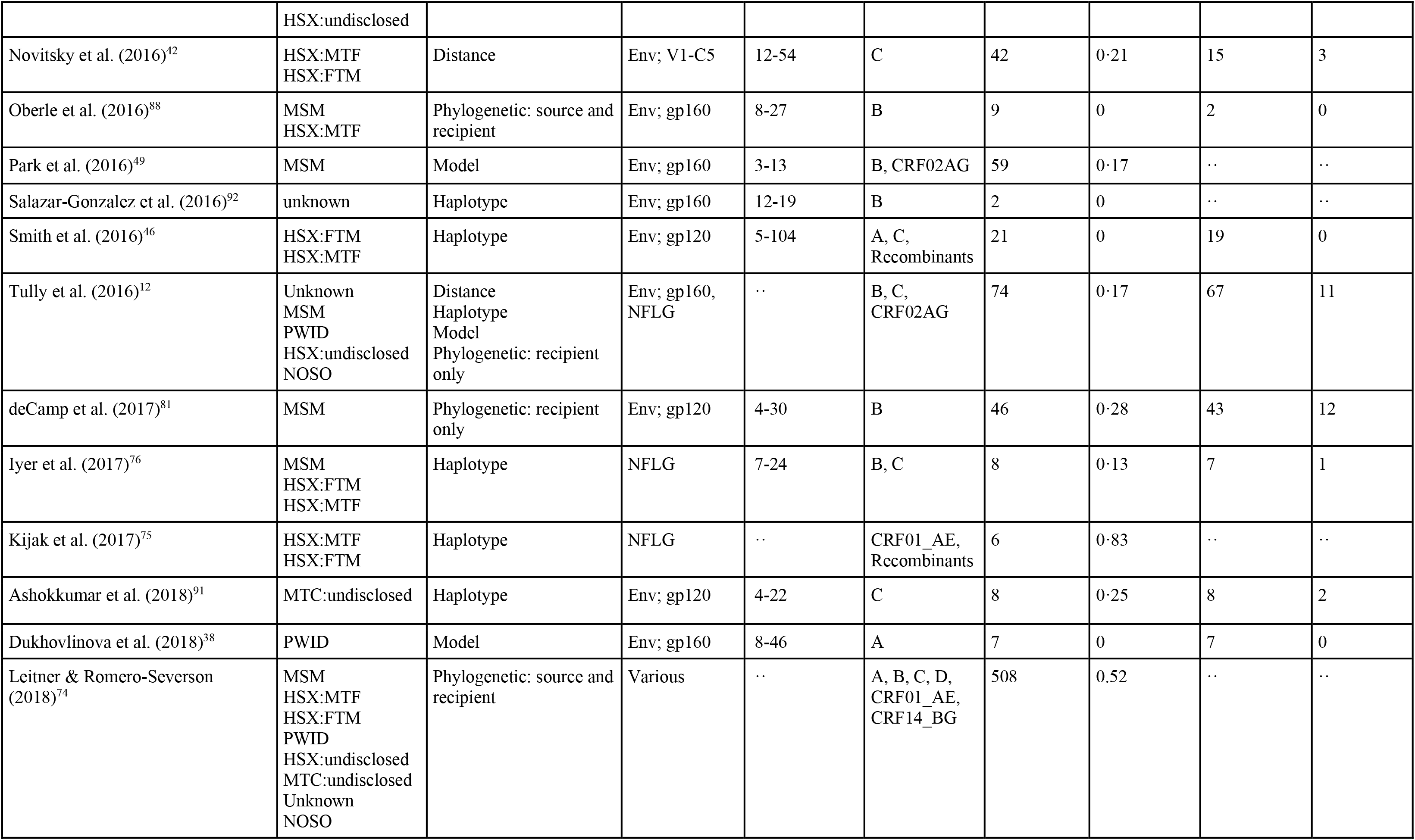
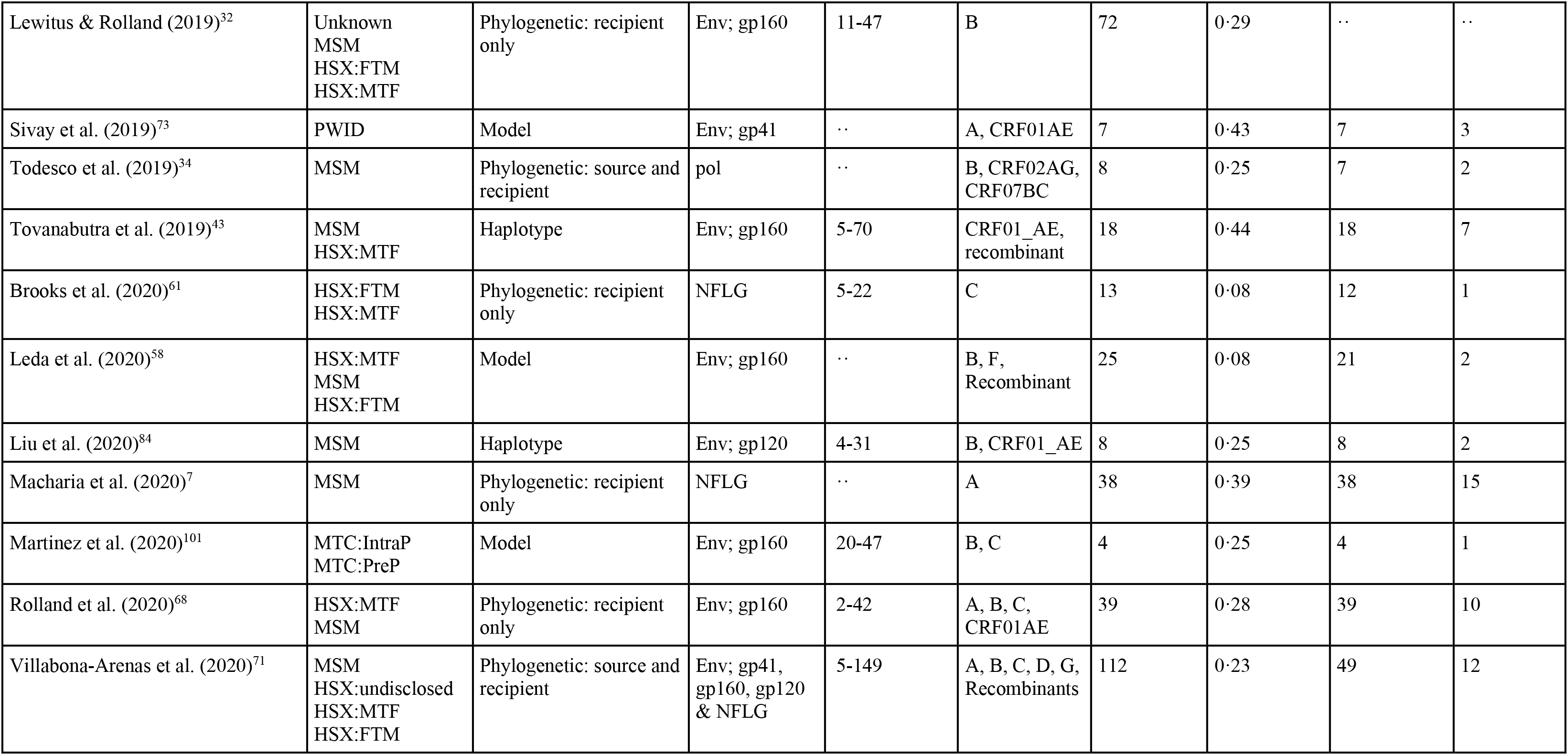
Included studies selected for inclusion from our systematic literature search. We record the route of transmission: female-to-male (HSX:FTM), male-to-female (HSX:MTF), men-who-have-sex-with-men (MSM), mother-to-child pre-partum (MTC:PreP), intrapartum (MTC:IntP) and post-partum (MTC:PostP); people who inject drugs (PWID), or nosocomial (NOSO). Additionally, we tabulate the method grouping used to infer founder multiplicity, the genomic region analysed, the number of participants analysed, and the proportion of infections initiated by multiple founders reported by each study. We note the number of single and multiple founder infections included within our base case dataset

|  | Transmission Routes | Method | Genomic Region | Range of Genomes Analysed Per Participant | Virus Subtype | Number of Participants | P(multiple founders) | Data Included |  |
| --- | --- | --- | --- | --- | --- | --- | --- | --- | --- |
|  |  |  |  |  |  |  |  | Participants | Multiple Founders |
| Wolinsky et al. (1992)<sup>79</sup> | MTC:undisclosed | Haplotype | Env; V3 & V4-V5 | 9-18 | Unknown | 3 | 0 | 3 | 0 |
| Briant et al. (1995)<sup>33</sup> | MTC:undisclosed | Phylogenetic: source and recipient | Env; V3 | .. | B | 4 | 0·75 | 4 | 3 |
| Poss et al. (1995)<sup>47</sup> | HSX:MTF | Haplotype | Env; gp120 | 10-17 | A, D | 6 | 0·83 | 6 | 5 |
| Wade et al. (1998)<sup>35</sup> | MTC:undisclosed<br>MTC:PreP | Phylogenetic: source and recipient | Gag; p17 | 49-115 | B | 2 | 0·5 | 2 | 1 |
| Long et al. (2000)<sup>52</sup> | HSX:MTF,<br>HSX:FTM | Molecular | Env; gp120 | .. | A, D, C,<br>Unknown | 36 | 0·55 | 36 | 15 |
| Dickover et al. (2001)<sup>72</sup> | MTC:IntraP<br>MTC:PreP | Molecular | Env; gp120 | .. | B | 23 | 0·26 | 23 | 6 |
| Delwart et al. (2002)<sup>63</sup> | HSX:FTM<br>HSX:MTF<br>Unknown | Molecular | Env; V3 | 2-141 | B | 17 | 0·06 | 17 | 1 |
| Learn et al. (2002)<sup>94</sup> | MSM | Molecular | Env; gp120 | .. | B | 8 | 0·5 | 8 | 4 |
| Long et al. (2002)<sup>96</sup> | HSX:MTF | Distance | Env; gp120 | .. | A, Unknown | 5 | 0·5 | 2 | 0 |
| Nowak et al. (2002)<sup>86</sup> | MTC:undisclosed | Phylogenetic: source and recipient | Env; V3 | 44-71 | B | 3 | 0·34 | 3 | 1 |
| Renjifo et al. (2003)<sup>64</sup> | MTC:PreP | Molecular | Env; gp120 | .. | A, C, D | 53 | 0·21 | 53 | 11 |
| Sagar et al. (2003)<sup>4</sup> | HSX:MTF | Molecular | Env; gp120 | .. | Unknown | 124 | 0·56 | 124 | 55 |
| Verhofstede et al. (2003)<sup>48</sup> | MTC:IntraP<br>MTC:PreP | Phylogenetic: source and recipient | Env; gp120 | 11-36 | A | 13 | 0·54 | 13 | 7 |
| Derdeyn et al. (2004)<sup>97</sup> | HSX:MTF<br>HSX:FTM | Phylogenetic: source and recipient | Env; gp120 | 13-20 | C, G | 7 | 0 | 7 | 0 |
| Ritola et al. (2004)<sup>70</sup> | HSX:MTF<br>HSX:FTM<br>MSM | Molecular | Env; V1-V3 | ·· | B | 26 | 0·52 | 25 | 7 |
| Sagar et al. (2004)<sup>29</sup> | HSX:MTF<br>PWID<br>MSM<br>HSX:FTM | Molecular | Env; V1-V5 | ·· | A, B,<br>Unknown | 17 | 0·24 | 17 | 4 |
| Sagar et al. (2006)<sup>60</sup> | HSX:MTF | Distance | Env; V1-V3 | 10-25 | A, D,<br>Unknown,<br>Recombinants | 12 | 0·5 | ·· | ·· |
| Gottlieb et al. (2008)<sup>62</sup> | MSM | Haplotype | Env; V1-V5 | 11-19 | B | 38 | 0·39 | 37 | 14 |
| Keele et al. (2008)<sup>3</sup> | PWID<br>MSM<br>Unknown<br>HSX:FTM<br>HSX:MTF | Distance<br>Haplotype<br>Model<br>Phylogenetic: recipient only | Env; gp160 | 10-67 | B | 102 | 0·24 | 44 | 15 |
| Kwiek et al. (2008)<sup>85</sup> | MTC:IntraP<br>MTC:PreP | Molecular | Env; V1-V2 | ·· | C | 48 | 0·42 | 48 | 28 |
| Salazar-Gonzalez et al. (2008)<sup>22</sup> | HSX:MTF<br>HSX:FTM | Distance<br>Haplotype<br>Phylogenetic: recipient only | Env; gp160 | 24-48 | C, Unknown | 12 | 0·34 | 12 | 4 |
| Abrahams et al. (2009)<sup>30</sup> | HSX:FTM<br>HSX:MTF | Distance<br>Model<br>Haplotype<br>Phylogenetic: recipient only | Env; gp160 | 15-42 | C, G | 69 | 0·22 | 69 | 15 |
| Haaland et al. (2009)<sup>14</sup> | HSX:MTF | Haplotype | Env; gp160 | 22-73 | A, C, | 27 | 0·23 | 22 | 3 |
|  | HSX:FTM | Phylogenetic: source and recipient |  |  | Unknown |  |  |  |  |
| Kearney et al. (2009)<sup>98</sup> | MSM<br>HSX:FTM<br>HSX:MTF<br>PWID | Phylogenetic: recipient only | pol | .. | B | 14 | 0·14 | 11 | 0 |
| Novitsky et al. (2009)<sup>51</sup> | HSX:MTF<br>HSX:FTM | Phylogenetic: recipient only | Env; gp120 | 11-33 | C | 8 | 0·25 | 8 | 2 |
| Salazar-Gonzalez et al. (2009)<sup>53</sup> | MSM<br>HSX:FTM | Distance<br>Haplotype<br>Model | NFLG | 4-26 | B, C | 12 | 0·083 | 2 | 0 |
| Bar et al. (2010)<sup>16</sup> | PWID | Phylogenetic: recipient only | Env; gp160 | 19-163 | B | 10 | 0·6 | 10 | 6 |
| Fischer et al. (2010)<sup>90</sup> | MSM | Model | Env; gp120 | .. | B | 3 | 0 | .. | .. |
| Li et al. (2010)<sup>15</sup> | MSM | Distance<br>Haplotype | Env; gp160 | 23-89 | B | 28 | 0·36 | 28 | 10 |
| Masharsky et al. (2010)<sup>17</sup> | PWID | Haplotype | env | 18-29 | A,<br>Recombinants | 13 | 0·31 | 13 | 4 |
| Zhang et al. (2010)<sup>77</sup> | MTC:IntraP | Phylogenetic: source and recipient | Env; V1-V5 | 25-30 | C,<br>Recombinants | 6 | 0 | 6 | 0 |
| Boeras et al. (2011)<sup>78</sup> | HSX:FTM<br>HSX:MTF | Phylogenetic: source and recipient | Env; V1-V4 | 31-73 | A, C | 8 | 0 | .. | .. |
| Collins-Fairclough et al. (2011)<sup>82</sup> | MSM<br>HSX:FTM<br>HSX:MTF<br>HSX:undisclosed | Haplotype | Env; V1-C4 | 5-20 | B | 27 | 0·23 | 14 | 2 |
| Herbeck et al. (2011)<sup>45</sup> | MSM | Distance | NFLG | 10-113 | B | 9 | 0·11 | 9 | 1 |
| Kishko et al. (2011)<sup>55</sup> | MTC:IntraP | Phylogenetic: source and recipient | Env; gp160 | 10-22 | B | 5 | 0·4 | 5 | 2 |
| Nofemela et al. (2011)<sup>44</sup> | HSX:MTF | Haplotype | env | 5-18 | A, B, C, D, | 22 | 0·27 | 22 | 6 |

**Table S3:** Included studies selected for inclusion from our systematic literature search. We record the route of transmission: female-to-male (HSX:FTM), male-to-female
|  |  |  |  |  | Recombinants |  |  |  |  |
| --- | --- | --- | --- | --- | --- | --- | --- | --- | --- |
| Novitsky et al. (2011)<sup>89</sup> | HSX:MTF<br>HSX:FTM | Distance<br>Haplotype<br>Model<br>Phylogenetic: recipient<br>only | gag & Env;<br>gp120 | 6-33 | C | 25 | 0·32 | 16 | 6 |
| Rachinger et al. (2011)<sup>87</sup> | MSM | Phylogenetic: source and<br>recipient | NFLG | ·· | B | 1 | 0 | ·· | ·· |
| Rieder et al. (2011)<sup>37</sup> | Unknown<br>MSM<br>HSX:MTF | Distance | Env; C2-V3-C3 | 14-16 | A, B, C, G,<br>CRF01AE,<br>CRF02AG,<br>CRF12BF,<br>CRF14BG | 143 | 0·11 | ·· | ·· |
| Rolland et al. (2011)<sup>54</sup> | MSM<br>HSX:MTF | Phylogenetic: recipient<br>only | NFLG | 2-14 | B, CRF02AG | 68 | 0·25 | 68 | 16 |
| Cornelissen et al. (2012)<sup>5</sup> | MSM | Phylogenetic: recipient<br>only | Env; V3-V4 | ·· | B | 31 | 0·13 | 31 | 4 |
| Henn et al. (2012)<sup>95</sup> | unknown | Distance | NFLG | ·· | B | 1 | 0 | ·· | ·· |
| Kiwelu et al. (2012)<sup>59</sup> | HSX:MTF | Phylogenetic: recipient<br>only | Env; gp120 | 5-62 | A, C, D | 50 | 0·27 | 43 | 10 |
| Rossenkhani et al. (2012)<sup>93</sup> | HSX:MTF<br>HSX:FTM | Phylogenetic: recipient<br>only | gag & Env;<br>gp120 | 3-92 | C | 20 | 0·15 | 5 | 0 |
| Sturdevant et al. (2012)<sup>36</sup> | MTC:undisclosed | Haplotype<br>Phylogenetic: recipient<br>only | Env; gp160 | 16-46 | C | 43 | 0·12 | 43 | 5 |
| Baalwa et al. (2013)<sup>69</sup> | HSX:MTF<br>HSX:FTM | Haplotype | NFLG | 20-82 | A, D,<br>Recombinants | 12 | 0·17 | 12 | 2 |
| Frange et al. (2013)<sup>80</sup> | MSM<br>HSX:MTF<br>HSX:FTM | Phylogenetic: source and<br>recipient | Env; C2-V5 | 19-43 | B | 8 | 0 | 8 | 0 |
| Chaillon et al. (2014)<sup>99</sup> | MTC:PreP | Phylogenetic: source and | Env; V1-V5 | 6-32 | CRF01_AE | 9 | 0·12 | 8 | 1 |
|  | MTC:IntraP | recipient |  |  |  |  |  |  |  |
| Sterrett et al. (2014)<sup>67</sup> | PWID | Distance<br>Haplotype<br>Model<br>Phylogenetic: recipient<br>only | Env; gp160 | 12-41 | B, CRF01AE,<br>CRF1501B,<br>Recombinants | 50 | 0·42 | 49 | 14 |
| Wagner et al. (2014)<sup>65</sup> | MSM<br>PWID | Phylogenetic: recipient<br>only | NFLG | ·· | B | 108 | 0·06 | 108 | 7 |
| Chen et al. (2015)<sup>41</sup> | MSM | Haplotype | Env; gp160 | 6-26 | B, CRF01AE,<br>CRF07BC | 30 | 0·2 | 18 | 3 |
| Danaviah et al.(2015)<sup>50</sup> | MTC:PostP | Phylogenetic: source and<br>recipient | Env; C2-V5 | ·· | C | 11 | 0·18 | 11 | 2 |
| Deymier et al. (2015)<sup>56</sup> | HSX:FTM | Phylogenetic: recipient<br>only | NFLG | 6-9 | C | 6 | 0 | 5 | 0 |
| Gounder et al. (2015)<sup>57</sup> | HSX:FTM<br>HSX:MTF | Phylogenetic: recipient<br>only | gag | 12-16 | C | 22 | 0·27 | 22 | 6 |
| Janes et al. (2015)<sup>6</sup> | MSM<br>HSX:FTM<br>HSX:MTF | Distance | Env; gp120 | 2-28 | B, CRF01AE | 163 | 0·29 | 100 | 32 |
| Le et al. (2015)<sup>102</sup> | PWID | Phylogenetic: source and<br>recipient | Env; gp120 | ·· | B | 2 | 0 | 2 | 0 |
| Zanini et al. (2015)<sup>100</sup> | HSX:MTF<br>MSM<br>HSX:FTM | Distance | NFLG | ·· | B, C,<br>CRF01AE | 9 | 0·22 | 9 | 2 |
| Chaillon et al. (2016)<sup>39</sup> | MSM<br>PWID | Distance<br>Phylogenetic: source and<br>recipient | Env; C2-V3 | ·· | B | 30 | 53·3 | 30 | 16 |
| Love et al. (2016)<sup>83</sup> | PWID<br>MSM<br>Unknown<br>HSX:FTM<br>HSX:MTF | Model | Env; gp160 | 10-163 | B, C | 182 | 0·23 | ·· | ·· |
|  | HSX:undisclosed |  |  |  |  |  |  |  |  |
| Novitsky et al. (2016)<sup>42</sup> | HSX:MTF<br>HSX:FTM | Distance | Env; V1-C5 | 12-54 | C | 42 | 0·21 | 15 | 3 |
| Oberle et al. (2016)<sup>88</sup> | MSM<br>HSX:MTF | Phylogenetic: source and recipient | Env; gp160 | 8-27 | B | 9 | 0 | 2 | 0 |
| Park et al. (2016)<sup>49</sup> | MSM | Model | Env; gp160 | 3-13 | B, CRF02AG | 59 | 0·17 | · | · |
| Salazar-Gonzalez et al. (2016)<sup>92</sup> | unknown | Haplotype | Env; gp160 | 12-19 | B | 2 | 0 | · | · |
| Smith et al. (2016)<sup>46</sup> | HSX:FTM<br>HSX:MTF | Haplotype | Env; gp120 | 5-104 | A, C,<br>Recombinants | 21 | 0 | 19 | 0 |
| Tully et al. (2016)<sup>12</sup> | Unknown<br>MSM<br>PWID<br>HSX:undisclosed<br>NOSO | Distance<br>Haplotype<br>Model<br>Phylogenetic: recipient only | Env; gp160,<br>NFLG | · | B, C,<br>CRF02AG | 74 | 0·17 | 67 | 11 |
| deCamp et al. (2017)<sup>81</sup> | MSM | Phylogenetic: recipient only | Env; gp120 | 4-30 | B | 46 | 0·28 | 43 | 12 |
| Iyer et al. (2017)<sup>76</sup> | MSM<br>HSX:FTM<br>HSX:MTF | Haplotype | NFLG | 7-24 | B, C | 8 | 0·13 | 7 | 1 |
| Kijak et al. (2017)<sup>75</sup> | HSX:MTF<br>HSX:FTM | Haplotype | NFLG | · | CRF01_AE,<br>Recombinants | 6 | 0·83 | · | · |
| Ashokkumar et al. (2018)<sup>91</sup> | MTC:undisclosed | Haplotype | Env; gp120 | 4-22 | C | 8 | 0·25 | 8 | 2 |
| Dukhovlinova et al. (2018)<sup>38</sup> | PWID | Model | Env; gp160 | 8-46 | A | 7 | 0 | 7 | 0 |
| Leitner & Romero-Severson (2018)<sup>74</sup> | MSM<br>HSX:MTF<br>HSX:FTM<br>PWID<br>HSX:undisclosed<br>MTC:undisclosed<br>Unknown<br>NOSO | Phylogenetic: source and recipient | Various | · | A, B, C, D,<br>CRF01_AE,<br>CRF14_BG | 508 | 0·52 | · | · |
| Lewitus & Rolland (2019)<sup>32</sup> | Unknown<br>MSM<br>HSX:FTM<br>HSX:MTF | Phylogenetic: recipient only | Env; gp160 | 11-47 | B | 72 | 0·29 | .. | .. |
| Sivay et al. (2019)<sup>73</sup> | PWID | Model | Env; gp41 | .. | A, CRF01AE | 7 | 0·43 | 7 | 3 |
| Todesco et al. (2019)<sup>34</sup> | MSM | Phylogenetic: source and recipient | pol | .. | B, CRF02AG, CRF07BC | 8 | 0·25 | 7 | 2 |
| Tovanabutra et al. (2019)<sup>43</sup> | MSM<br>HSX:MTF | Haplotype | Env; gp160 | 5-70 | CRF01_AE, recombinant | 18 | 0·44 | 18 | 7 |
| Brooks et al. (2020)<sup>61</sup> | HSX:FTM<br>HSX:MTF | Phylogenetic: recipient only | NFLG | 5-22 | C | 13 | 0·08 | 12 | 1 |
| Leda et al. (2020)<sup>58</sup> | HSX:MTF<br>MSM<br>HSX:FTM | Model | Env; gp160 | .. | B, F, Recombinant | 25 | 0·08 | 21 | 2 |
| Liu et al. (2020)<sup>84</sup> | MSM | Haplotype | Env; gp120 | 4-31 | B, CRF01_AE | 8 | 0·25 | 8 | 2 |
| Macharia et al. (2020)<sup>7</sup> | MSM | Phylogenetic: recipient only | NFLG | .. | A | 38 | 0·39 | 38 | 15 |
| Martinez et al. (2020)<sup>101</sup> | MTC:IntraP<br>MTC:PreP | Model | Env; gp160 | 20-47 | B, C | 4 | 0·25 | 4 | 1 |
| Rolland et al. (2020)<sup>68</sup> | HSX:MTF<br>MSM | Phylogenetic: recipient only | Env; gp160 | 2-42 | A, B, C, CRF01AE | 39 | 0·28 | 39 | 10 |
| Villabona-Arenas et al. (2020)<sup>71</sup> | MSM<br>HSX:undisclosed<br>HSX:MTF<br>HSX:FTM | Phylogenetic: source and recipient | Env; gp41, gp160, gp120 & NFLG | 5-149 | A, B, C, D, G, Recombinants | 112 | 0·23 | 49 | 12 |

### Temporal Structure of Exposure and Method

**Figure S1:**
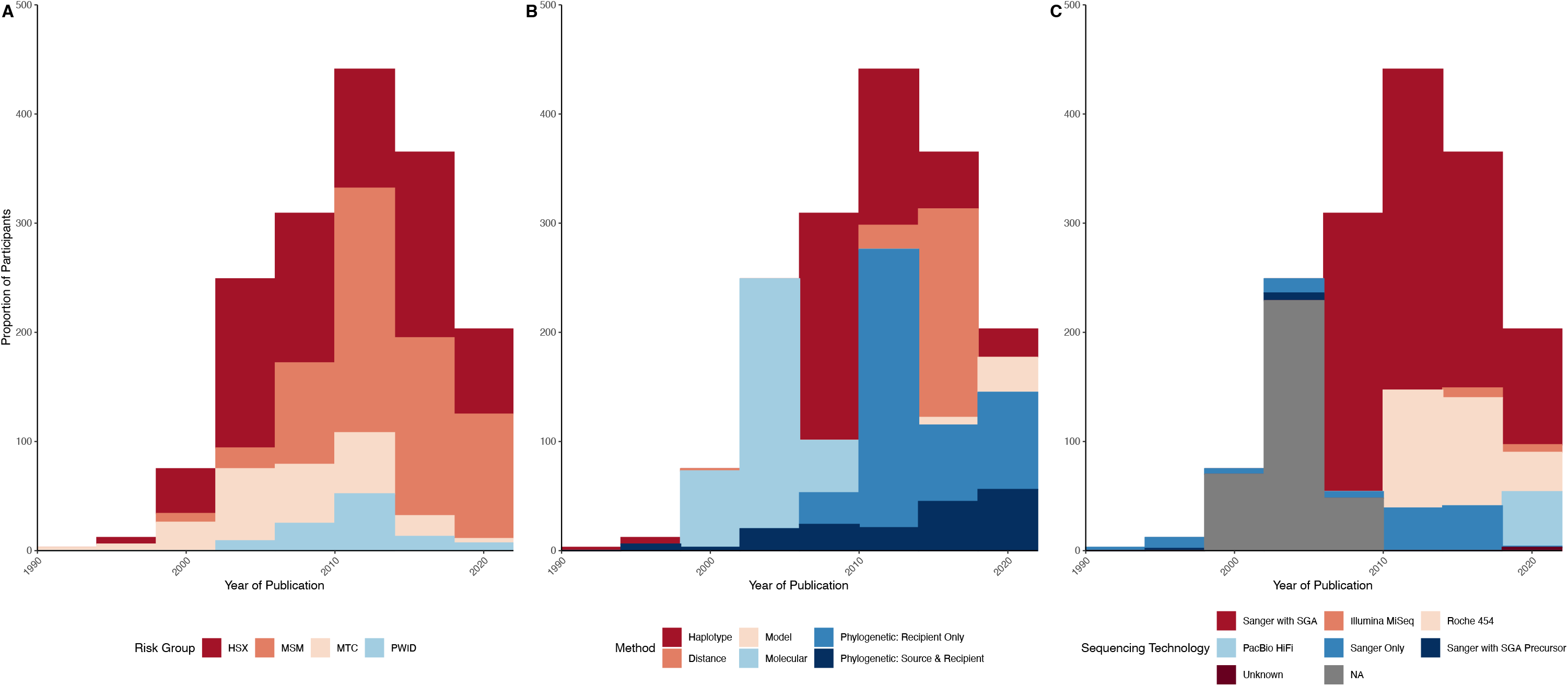
Distributions of transmission route (A), grouped method (B) and sequencing technology (C) over time, highlighting the epidemiologic and methodological step-changes that occurred over the three decades in which the selected studies were published. This means that earlier methods may be biased to those transmission routes that were more common in earlier studies.

### Sensitivity Analyses of Pooling

**Figure S2:**
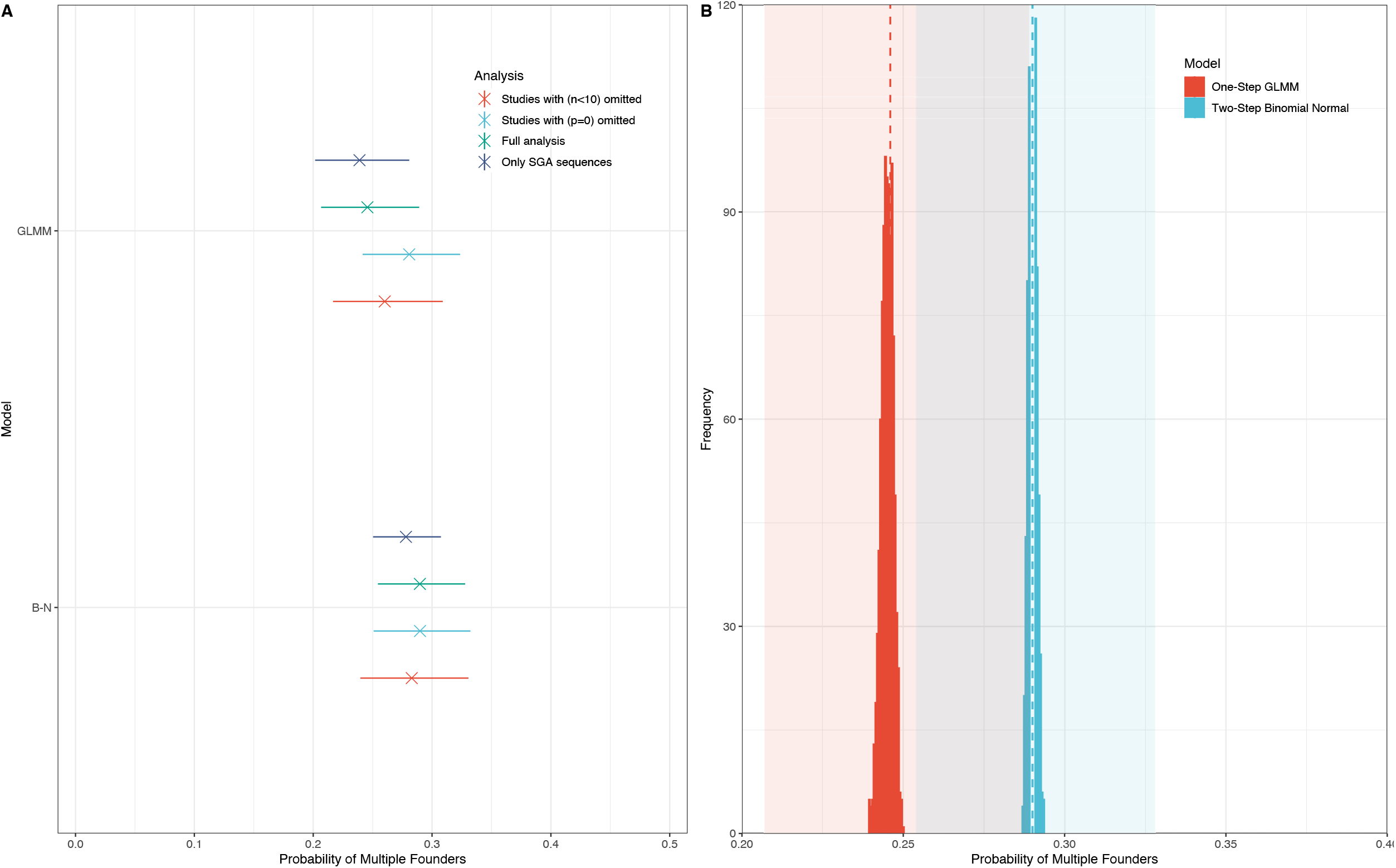
A comparison of the pooled estimates of the probability that an infection is initiated by multiple founders by the one-step (GLMM) and two-step (Binomial-Normal (B-N)) models and respective sensitivity analyses. Plot (A) shows both models calculate concordant estimates and are robust to sensitivity analyses designed to test our inclusion/exclusion criteria, and biases introduced by small or minimal-effect studies. B) reports the distribution of estimates, recalculated from 1000 datasets in which the representative datapoint for each individual was sampled at random from a pool of their possible measurements. The dashed lines and shaded areas denote the original point estimate and confidence intervals, respectively.

### Influence of Methodology and Number of Genomes Analysed

We analysed data from participants spanning the interquartile range (11 - 28 genomes), and then restricted the analysis to participants with higher than the upper quartile value (numbers of genomes>28) or lower than the lower quartile value (number of genomes <11). Restricting the analysis to participants for whom a large (>28) or small (<11) number of sequences were analysed adjusted the pooled estimate to 0·26 (0·20-0·34) and 0·21 (0·17-0·25), respectively (Fig.S3). The model fitted to participants spanning the interquartile range also revealed a slight increase in the probability of observing multiple founder variants when compared to the original estimates (0·27 (0·24-0·31)). These findings suggest the presence of a subtle correlation between the number of genomes analysed and the probability of observing multiple founder variants.

**Figure S3:**
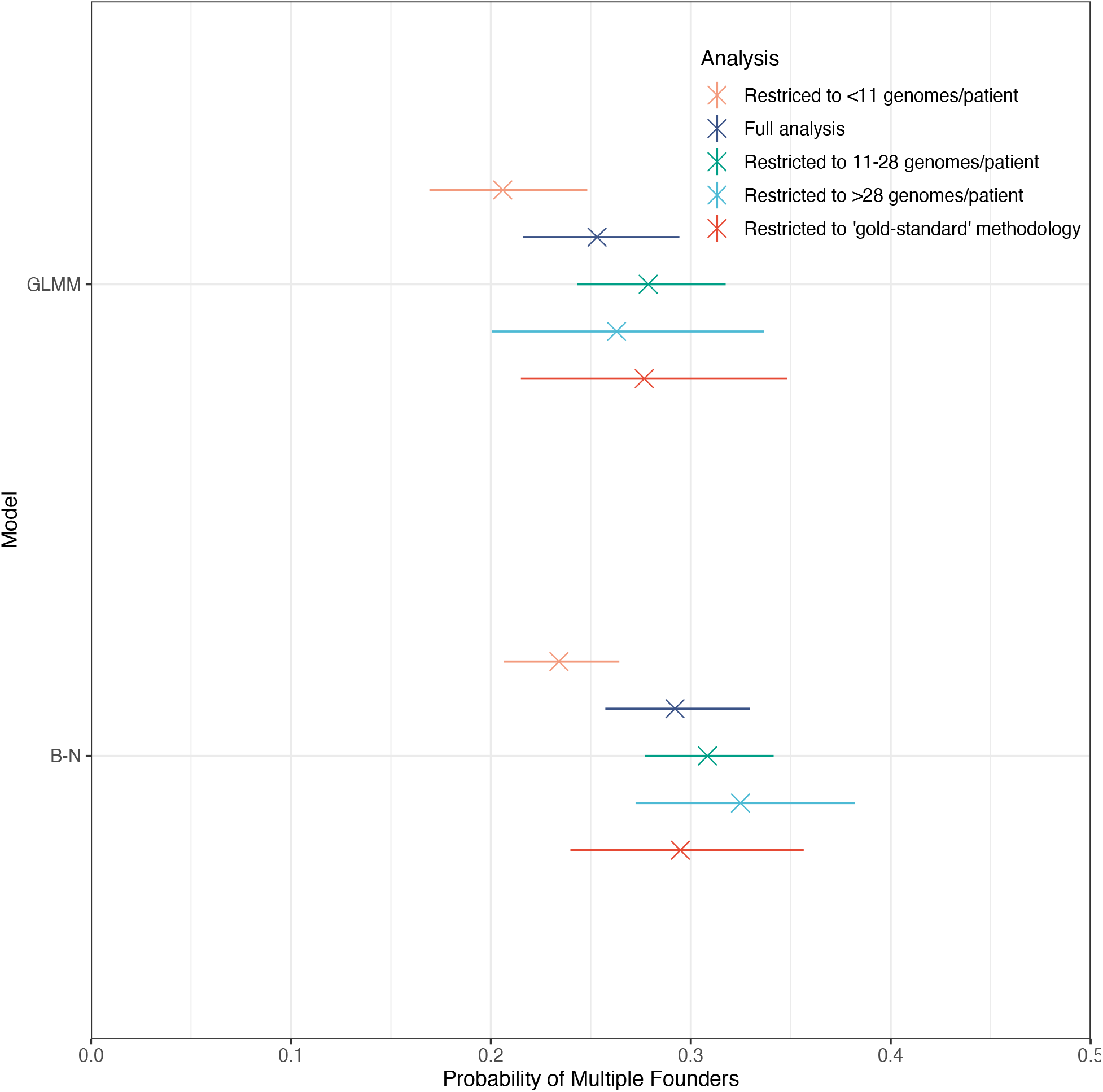
Comparing the pooled estimates of the probability that an infection is initiated by multiple founders by the one-step (GLMM) and two-step (Binomial-Normal (B-N)) models under our ‘gold-standard’ methodology, and when varying the threshold of the number of genomes analysed per patient.

### Leave-One-Out Cross Validation: Studies

**Figure S4:**
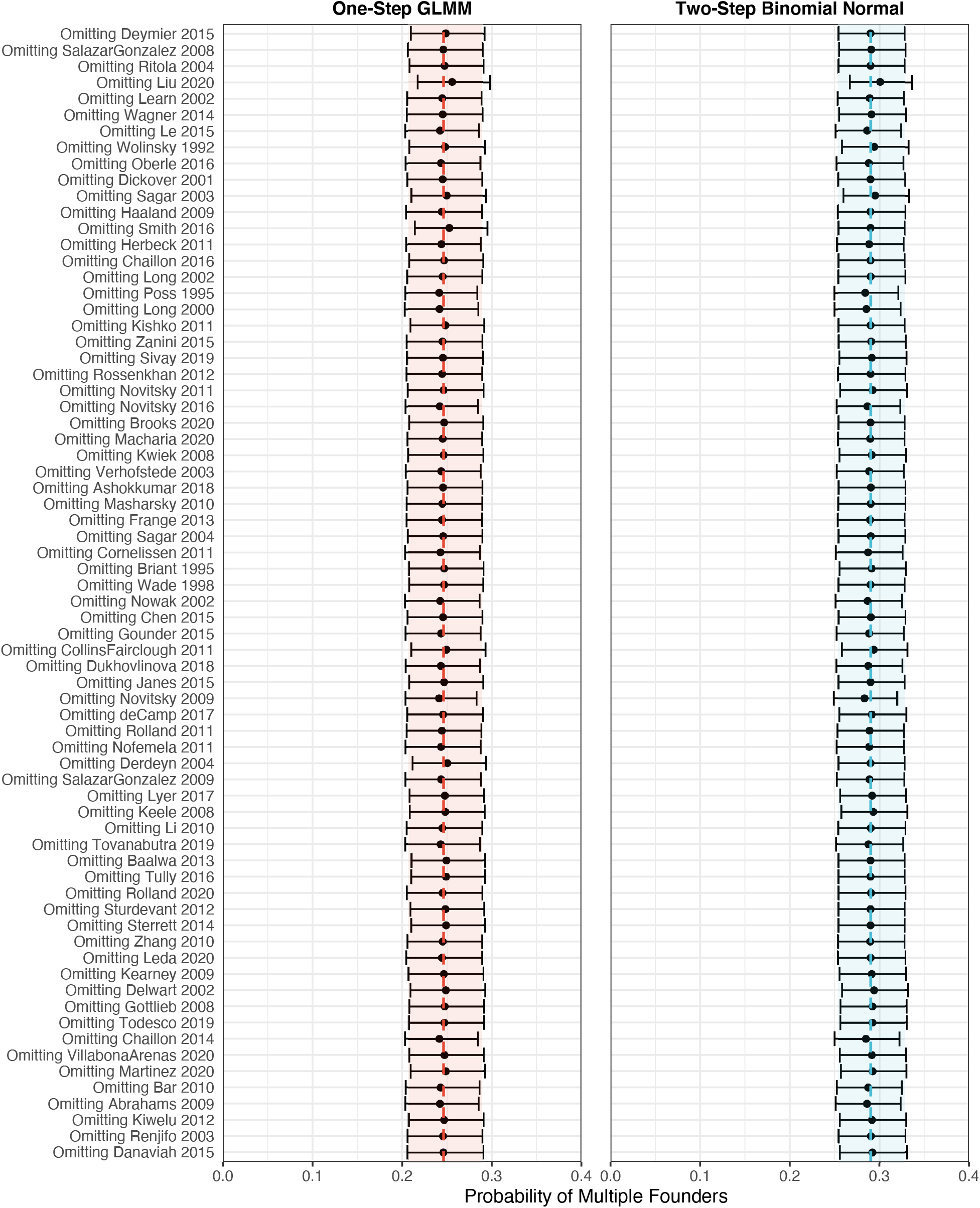
For both one-step and two-step models, we visually inspect the influence of each study included in our analysis on the pooled estimate that an infection is initiated by multiple founders. We find that in iteratively excluding individual studies, no discernible impact on the overall pooled estimate is made. The dashed lines and shaded areas denote the original point estimate and confidence intervals, respectively.

### Leave-One-Out Cross Validation: Transmission Routes

**Figure S5:**
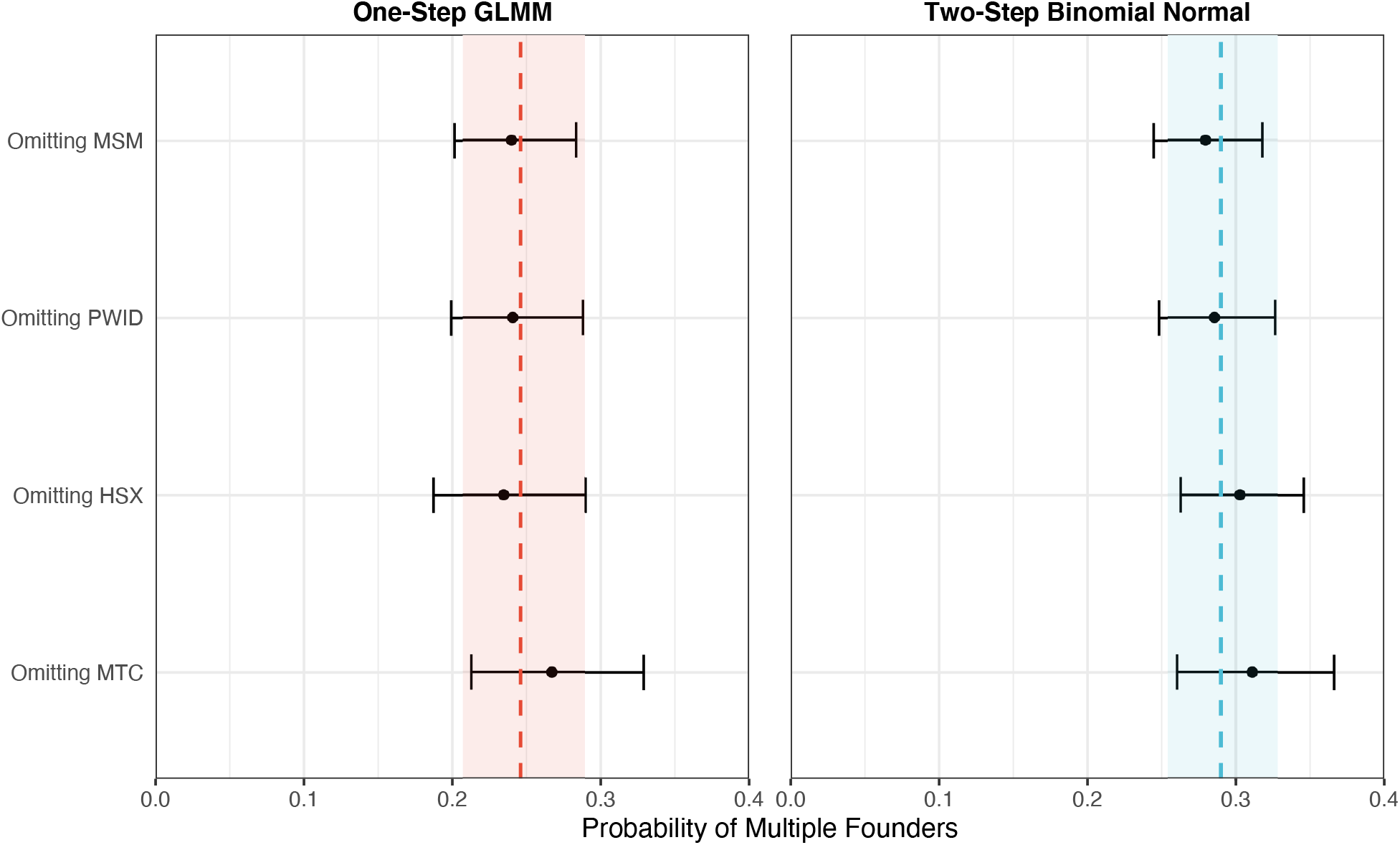
For both one-step and two-step models, we visually inspect the influence of each risk group included in our analysis on the pooled estimate that an infection is initiated by multiple founders. We find no discernible impact on the overall pooled estimate is made. The dashed lines and shaded areas denote the original point estimate and confidence intervals, respectively.

### Comparison of Vaccine Escape and Placebo Participants

Some of the selected studies included participants enrolled on vaccine trials. As breakthrough infections of vaccine-recipients may not reflect natural infection, we compare vaccine and placebo arms of trials for which these data were available. This analysis included participants from HTVN502 and RV144 (A third vaccine trial (HTVN505) is not included as participant vaccine status was not available). Estimates of founder multiplicity were extracted from Rolland et al 2011 (HTVN502), and Janes et al 2015 (RV144), following our inclusion criteria of selecting the first instance for which data are available (HTVN502 participants were also subsequently analysed by Janes et al.). We did not find any significant difference between vaccine-breakthrough and placebo infections.

**Figure S6:**
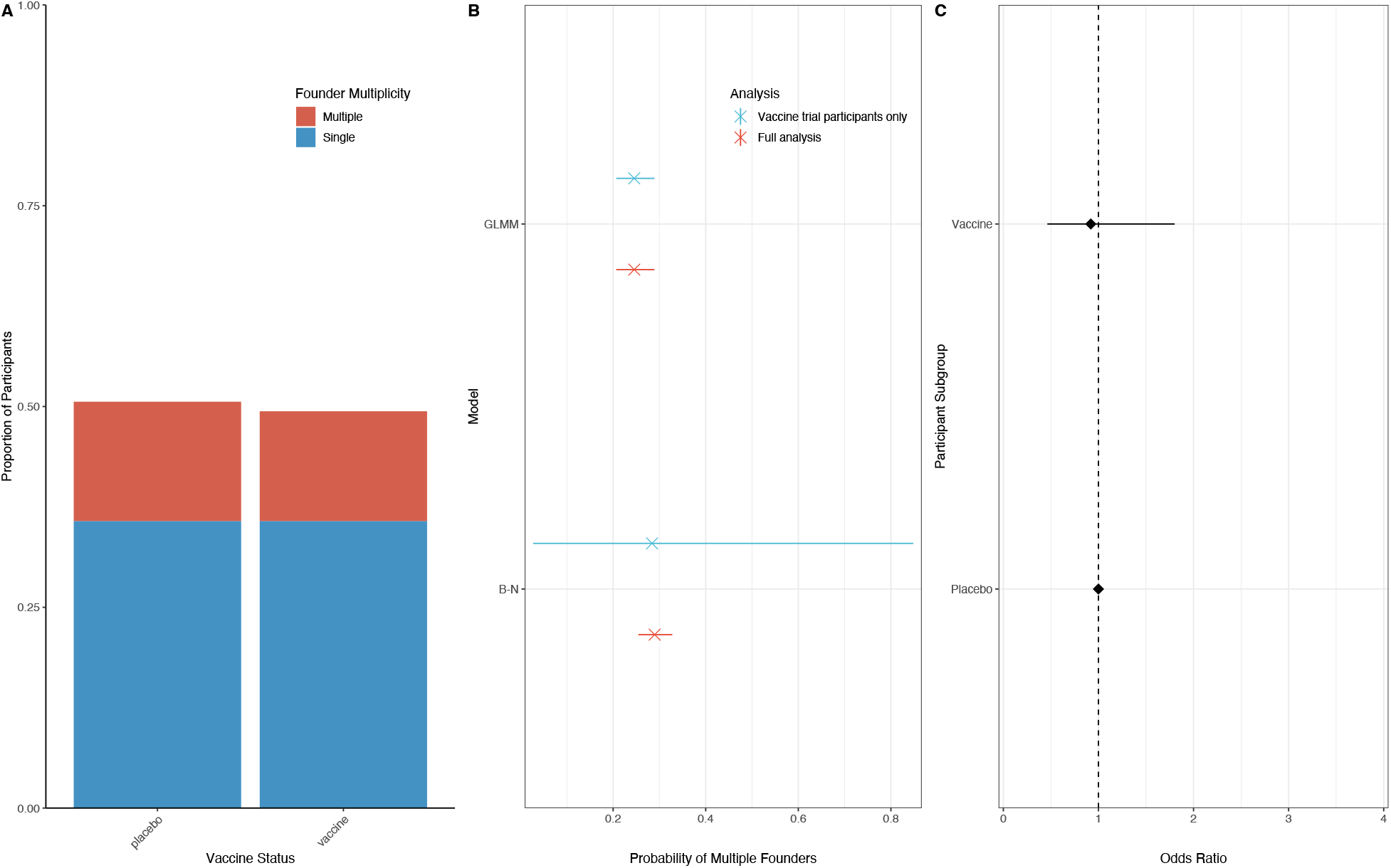
A) The proportion of infections identified as being initiated by multiple founders, segregated by vaccine status. B) Pooling estimates calculated using one and two-step models for vaccine trial only datapoints, compared to the base case dataset. C) Univariable analysis finding no significant difference in the odds of observing multiple founder variants between vaccinated and placebo arms of vaccine trial participants.

### Comparison of Sequencing Technologies

Of 70 selected studies in our base case dataset, eleven studies used deep sequencing (Roche 454 – 9 studies, Ilumina – 1 study, PacBio HiFi-1 study). To investigate whether the higher resolution of deep sequencing approaches influenced the observation of multiple founder variants initiating HIV infection, we conducted a univariable regression across those studies that used sequence-based methods. (0.22 (95% CI: 0.19-0.27)) was slightly lower than our original pooled estimate (0·25 (95% CI: 0·21-0·29)). In our univariable analysis, we did not find the odds of observing multiple founder variants differed significantly across sequencing methodologies.

**Figure S7:**
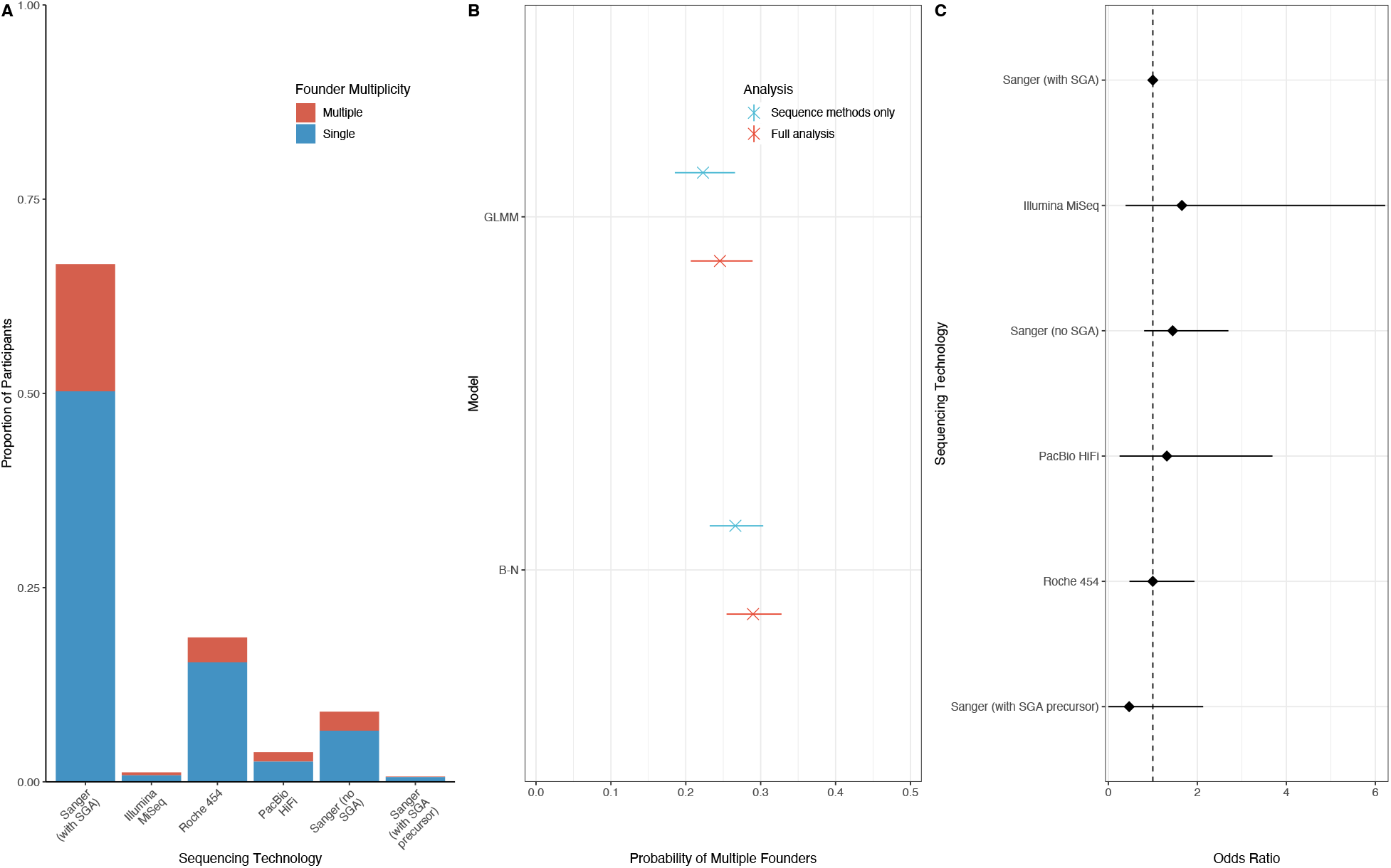
A) The proportion of infections identified as being initiated by multiple founders, segregated by sequencing technology. B) Pooling estimates calculated using one and two-step models for sequence methods only datapoints, compared to the base case dataset. C) Univariable analysis finding no significant difference in the odds of observing multiple founder variants across sequencing technologies.

### Evaluating the Impact of Molecular Methods

Both our multivariable and univariable analyses identified differences in the probability of multiple founders according to the genomic region analysed. Molecular methods, which we defined as approaches that rely on the formation of heteroduplexes during gel electrophoresis of viral RNA, are very sensitive. This allows one to distinguish genetically similar and dissimilar segments, however the use of these methods on short fragments of envelope may produce false positive results. Indeed, in both our univariable and multivariable analyses, there were significantly greater odds of recording multiple founder infections if molecular methods were used. To evaluate the impact of molecular methods as a confounder on the genomic region, we recalculated our pooled estimate under different scenarios and re-fitted a univariable model of genomic region in the absence of molecular methods. Of 1657 individuals, 1315 in our base case dataset were analysed using the envelope genomic region. Pooled estimates for envelope only individuals and envelope only individuals without molecular methods under the GLMM were 0·28 (0·23-0·32) and (0·25 0·21-0·29) respectively. A univariable analysis of genomic region fitted to the main dataset excluding molecular methods reported findings consistent with the main univariable analysis.

**Figure S8:**
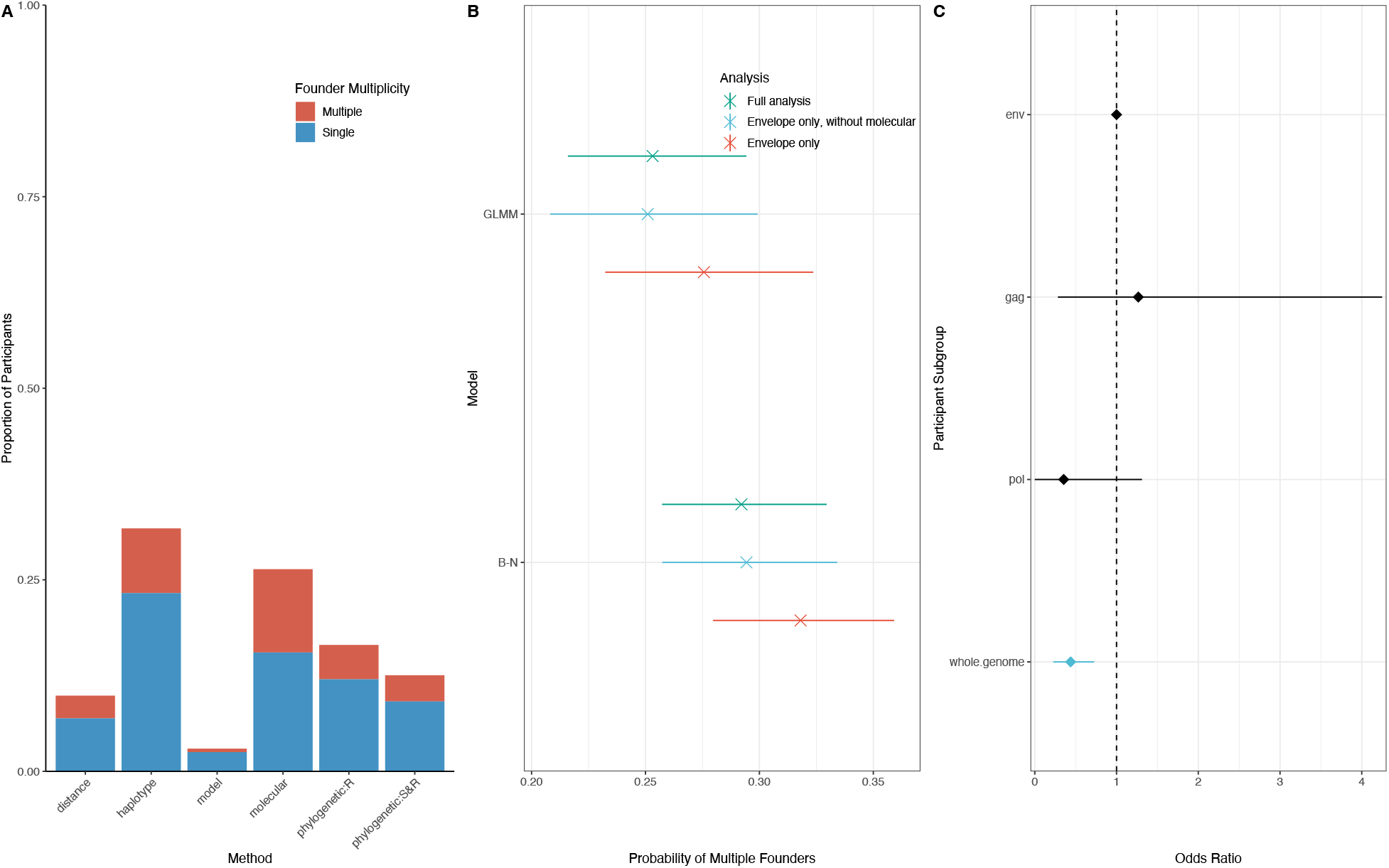
A) The founder variant multiplicity of 1315 individuals was analysed using the envelope genomic region, here segregated by method and indicating the prevalence of multiple founder infections. B) Pooled estimates calculated using one and two-step models from individuals for whom the envelope genomic region was analysed including/excluding molecular methods. C) Univariable analysis on dataset excluding molecular methods, reporting findings consistent with the main univariable analysis.

### Evaluation of Publication Bias

**Figure S9:**
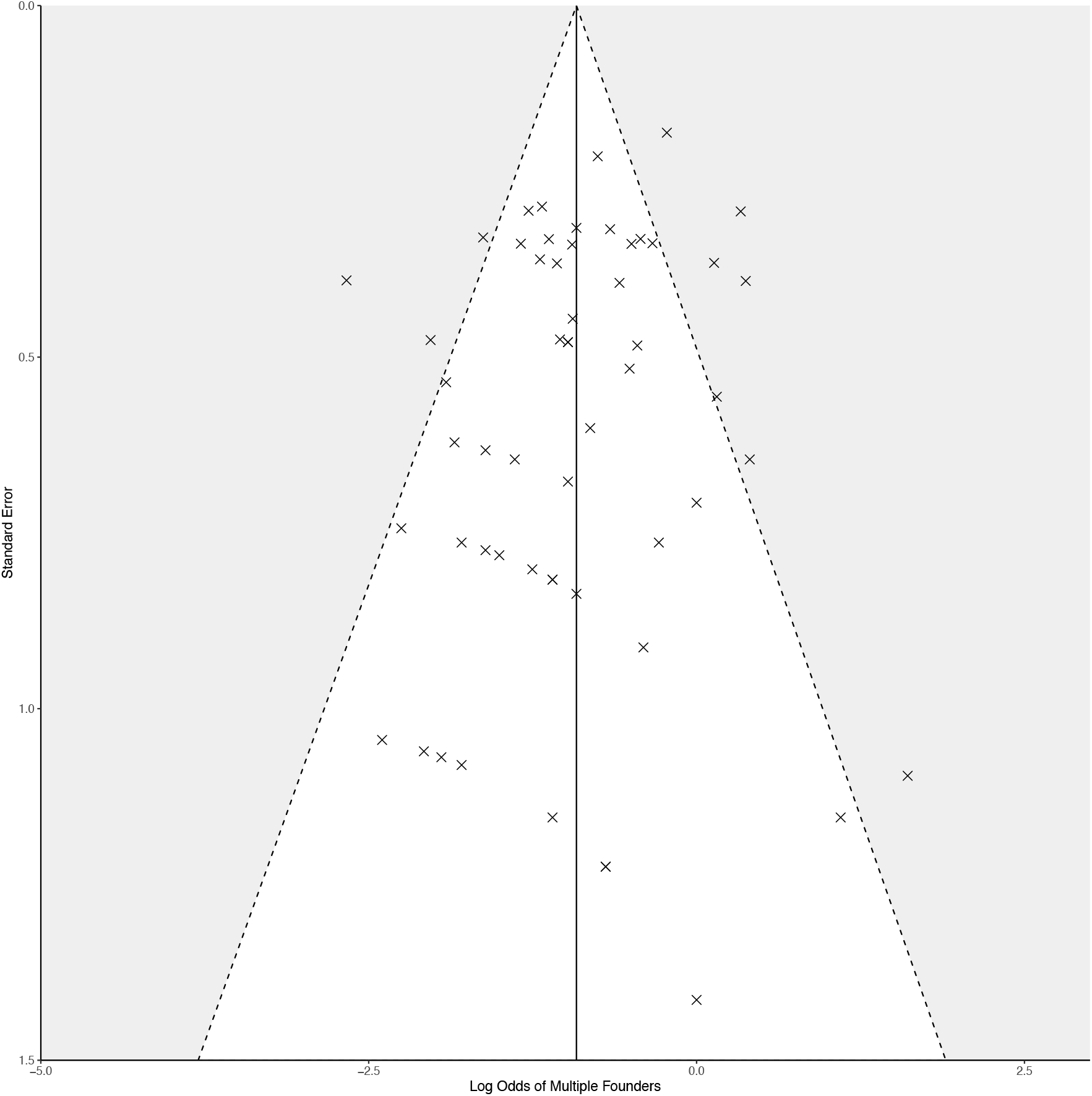
Funnel plot to visually evaluate the presence of publication bias. In the absence of publication bias, study estimates are distributed symmetrically with respect to the pooled estimate (vertical solid black line). Here, the log odds of an infection being initiated by multiple founders for each study, plotted against the standard error for each study indicate an absence of publication bias. This conclusion was supported by an Egger’s Regression Test: t = - 0·7495, df = 55, p = 0·4568.

### Binned Residuals Plot

**Figure S10:**
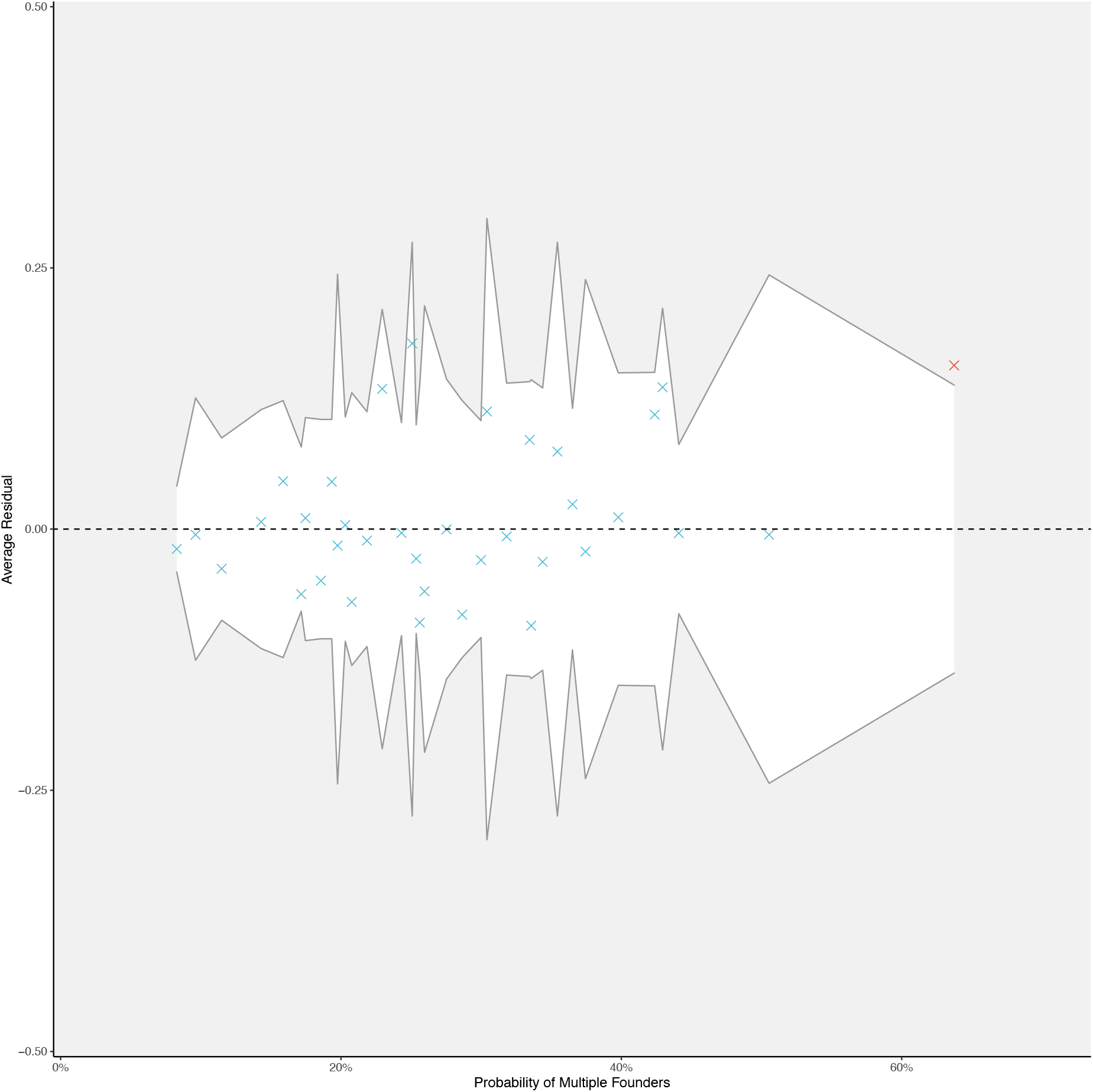
Binned residuals from the select multivariable model. 97% of the average residuals across each bin fall within the 95% confidence intervals (white area), indicating a good model fit.

### Sensitivity Analyses for Meta-regression

**Figure S11:**
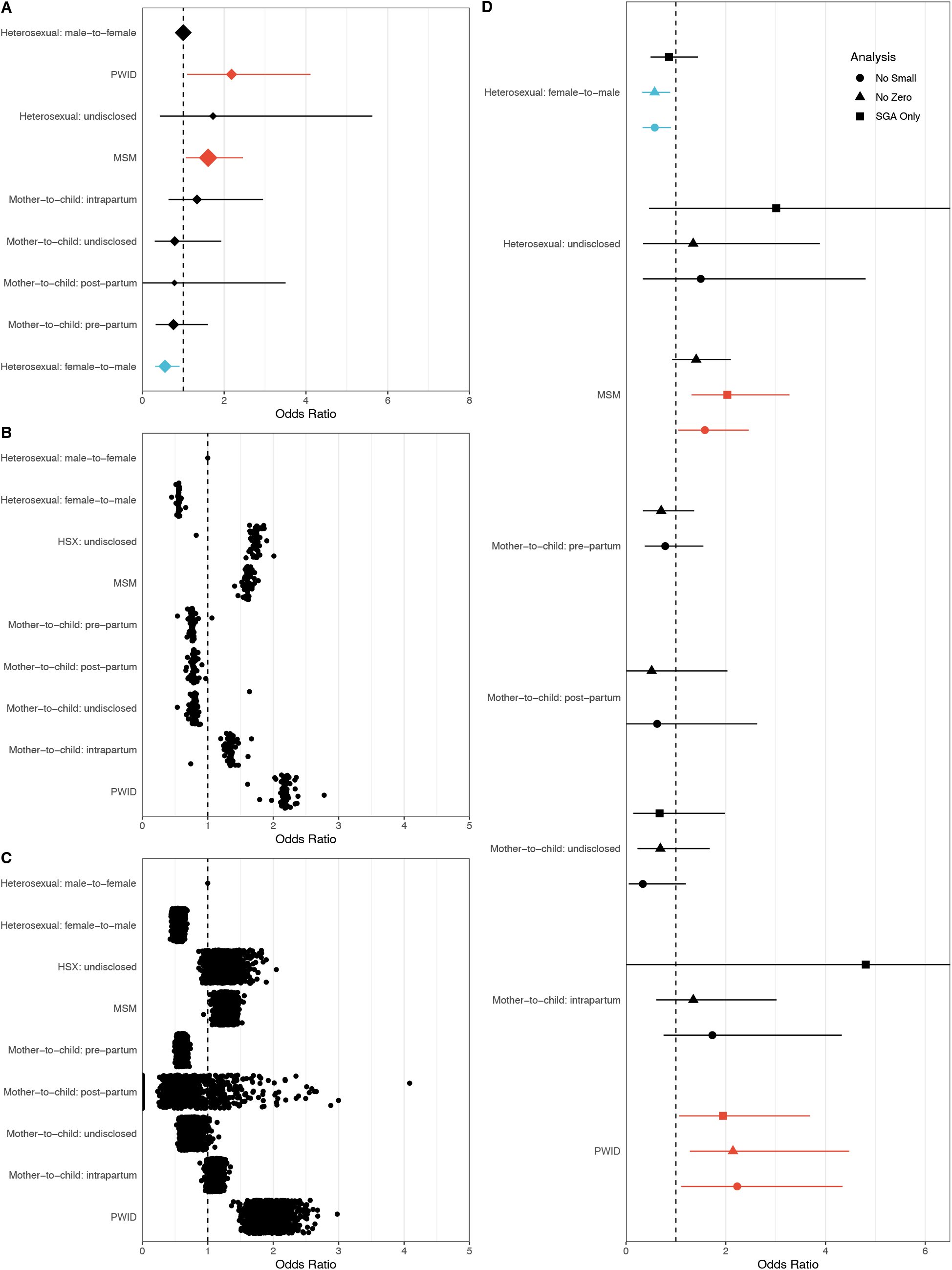
Odds ratios that an infection is initiated by multiple founders, stratified by route of transmission, as calculated in the main analysis (A), following the iterative exclusion of individual studies (B) and bootstrapped estimates recalculated from 1000 datasets in which the representative datapoint for each individual was sampled at random from a pool of their possible measurements (C). Panel (D) plots the odds ratios of all covariate levels included in the meta-regression, stratifying by previously defined sensitivity analyses. Overly generous confidence intervals in (D), particularly under the condition of single genome analysis (SGA) only data, is likely due to small sample sizes in at those levels (n<10).

